# Alpha-synuclein propagation trajectories in a population-based postmortem cohort

**DOI:** 10.64898/2026.01.13.26343943

**Authors:** Parsa Khalafi, Jacob W Vogel, Per Boghammer, Anna Raunio, Ville Kivistö, Sara Savola, Eloise H Kok, Elizaveta Mikhailenko, Liisa Myllykangas, Alain Dagher

## Abstract

Parkinson’s disease and dementia with Lewy bodies are defined neuropathologically by the accumulation of α-synuclein. Postmortem studies have led to theories that pathology propagates systematically along anatomical brain networks. Yet, existing staging systems present conflicting patterns of pathology distribution. Recent work suggests that these discrepancies may reflect heterogeneous “brain-first” and “body-first” subtypes, but prior studies are largely based on clinical cohorts that underrepresent incidental pathology in the general aging population. Previous studies have also rarely examined spinal cord involvement, useful for distinguishing routes of propagation from the body. Using data-driven disease-progression modelling in a population-based cohort of 304 individuals aged 85 years and older, we identify three distinct α-synuclein propagation trajectories. We find a “body-first” subtype (26.8%) originating in the lower brainstem, and two divergent “brain-first” subtypes (73.2%) initiating in the olfactory bulb, one spreading toward the brainstem and the other preferentially targeting the amygdala. The amygdala-directed trajectory is strongly associated with Alzheimer’s co-pathology, APOE ε4 genotype, and poorer cognitive outcomes. These findings reconcile discrepancies among prior staging frameworks by demonstrating substantial spatial heterogeneity even within brain-originating pathways, and they establish the biological distinctness of these trajectories in an unselected, community-dwelling population. Moreover, we show that the spatial extent of α-synuclein pathology contributes to cognitive impairment independently of comorbid Alzheimer’s disease or transactive response DNA-binding protein 43 (TDP-43) pathology. Together, these results provide a framework for patient stratification and for linking α-synuclein propagation patterns to clinical trajectories from prodromal to late stages.

**Abbreviated Summary:** Using data-driven modelling in a population-based cohort (Vantaa 85+), Khalafi et al. identify three distinct α-synuclein propagation trajectories: one originating in the lower brainstem and two initiating in the olfactory bulb with divergent downstream targets. The trajectory preferentially involving the amygdala is strongly associated with Alzheimer’s co-pathology and cognitive decline, independent of other comorbid pathologies.

## Introduction

Parkinson’s disease and dementia with Lewy bodies are characterized by the aberrant aggregation and propagation of alpha-synuclein (αSyn), forming Lewy bodies and Lewy neurites in neurons.^1,2^ These disorders represent the second most common neurodegenerative pathology after Alzheimer’s disease, affecting up to 10% of individuals over 75 years old.^2–4^ Incidental Lewy body disease, defined by αSyn pathology in the absence of overt clinical symptoms, is notably prevalent, occurring in up to 30% of individuals over the age of 65.^5^ The complexity of synucleinopathies is further heightened by their frequent co-occurrence with Alzheimer’s disease-related pathologies, including amyloid-*β* (A*β*) plaques and tau neurofibrillary tangles (NFTs), which confound clinical phenotypes and accelerate cognitive decline.

Neuropathological observations have yielded crucial insights into the neurodegenerative process in Parkinson’s disease. Braak and colleagues, through systematic postmortem analyses, suggested that Parkinson’s disease pathology follows a stereotyped and progressive pattern of propagation through the nervous system.^6,7^ They defined a series of sequential stages of pathology progression, each associated with characteristic clinical manifestations. The observation that the olfactory bulb and lower brainstem [dorsal motor nucleus of the vagus (DMV)] are among the earliest affected regions suggested that the pathological process may originate in peripheral sites and propagate in a caudo-rostral trajectory from the lower brainstem to limbic and neocortical regions. More recently, animal models have demonstrated that misfolded αSyn isoforms can propagate in a prion-like manner across interconnected brain regions, providing further mechanistic support for the Braak hypothesis.^8,9^

In parallel, alternative staging systems based on postmortem analyses have been proposed. The Dementia with Lewy Bodies Consortium (DLBC) consensus guidelines emphasize a similar caudo-rostral spread of pathology in dementia with Lewy bodies, with early involvement of the brainstem.^10–12^ The Unified Staging System for Lewy Body Disorders (USSLB) integrates these models for Parkinson’s disease and dementia with Lewy bodies, as well as incidental Lewy body disease. Unlike previous systems, it suggests that pathology begins in the olfactory bulb, later progressing into brainstem-predominant or limbic-predominant pathways before equally involving both regions and finally reaching the neocortex.^13^

More recently, the α-Synuclein Origin Site and Connectome (SOC) model was introduced to account for clinical and biological heterogeneity by proposing distinct disease epicentres.^14,15^ Grounded in animal models and human computational studies,^8,16–19^ this framework suggests that pathology propagates via the connectome from two potential initiation points: a ‘body–first’ pathway originating in the enteric nervous system and spreading caudo–rostrally, or a ‘brain–first’ pathway beginning in the olfactory bulb or amygdala and progressing rostro–caudally^20^ —each linked to specific clinical patterns. Resolving these divergent frameworks is therefore critical to characterize individual disease trajectories, map neuropathological progression to clinical milestones, and elucidate the biological underpinnings of clinical heterogeneity.

Despite these advances, consensus on these staging systems remains elusive. Current frameworks are primarily derived from clinically ascertained cohorts, which disproportionately represent advanced disease stages. Incidental Lewy body cases, thought to represent prodromal forms of Parkinson’s disease or dementia with Lewy bodies, are rarely included in these datasets, despite likely reflecting the earliest phases of αSyn progression. The initial accumulation of αSyn is believed to occur years before the onset of clinical symptoms,^14,21^ and this prolonged latency represents a critical yet underexplored window for early-stage investigation. Moreover, these frameworks often assume an overly stereotyped progression, failing to accommodate inter-individual variability or to quantify uncertainty in disease trajectories. Data-driven approaches offer a promising avenue to resolve these limitations.^22^ The Subtype and Stage Inference (SuStaIn) algorithm, an extension of event-based modeling, reconstructs probabilistic temporal sequences of pathological events, identifying latent subtypes while accounting for variability in regional involvement.^23^ Prior applications of SuStaIn restricted to hospital-based autopsy cohorts have revealed multiple αSyn progression patterns, including trajectories initiating in the olfactory system, brainstem, and sympathetic trunk.^24,25^ However, these studies were limited by their focus on clinically diagnosed cases in which pathology is advanced and widespread. Population-based cohorts, which encompass the full spectrum of aging-related pathology, including early and incidental forms, are essential for validating and refining these models.^26^ In addition, the integration of spinal cord samples may further help to elucidate the interplay between central and peripheral pathways.^26–28^

Here, we applied the ordinal adaptation of SuStaIn to postmortem neuropathological data from the Vantaa 85+ study, a prospective, population-based cohort of 304 individuals aged ≥85 years at baseline, with comprehensive sampling across 19 brain, spinal cord, and peripheral regions. Among 139 subjects with detectable αSyn pathology, we identified three distinct progression subtypes originating in either olfactory or lower brainstem regions, revealing divergent brain-first and body-first pathways. These subtypes aligned with established staging systems but provided enhanced resolution of spatial heterogeneity, associating specific trajectories with Alzheimer’s disease co-pathology, apolipoprotein E (APOE) ε4 genotype, dementia with Lewy bodies likelihood, and cognitive outcomes.

## Materials and methods

### Study population

The Vantaa 85+ study cohort includes all individuals aged 85 years or older in the city of Vantaa (Southern Finland) on April 1, 1991 (*n*=601).^26,29–33^ Of them, 565 eligible subjects died during the 10-year follow-up, and postmortem neuropathological autopsies were performed in 304. Post-mortem examinations were not determined by the cause of death, and comparisons between the neuropathologically examined subgroup and the overall study population revealed no significant differences in age at death or sex.^26^ This prospective, population-based study received ethical approval from both the Health Centre of Vantaa and Helsinki University Hospital, and all participants and/or their relatives provided informed consent for the Vantaa 85+ study, and the relatives also gave written consent for autopsy.

### Clinical and genetic assessment

Global cognitive function was assessed using the Mini-Mental State Examination (MMSE) recorded closest to the time of death.^34^ Cognitive status was classified as unimpaired (MMSE 26–30), mildly impaired (MMSE 21–25), moderately impaired (MMSE 11–20), or severely impaired (MMSE 0–10). Dementia was diagnosed if subjects met the criteria specified in the Revised Third Edition of the Diagnostic and Statistical Manual of Mental Disorders (DSM-III-R) and had experienced symptoms for at least three months. This diagnosis was confirmed by consensus between two neurologists, with the duration of dementia defined as the interval from diagnosis to death.^30,31^ APOE genotyping was performed as previously described.^35^

### Neuropathological assessment

During autopsy, standard tissue samples were obtained from at least 16 brain regions, 4 spinal cord regions (from 4 different spinal cord levels), and 2 peripheral regions (see below). Samples from any damaged or missing areas were excluded. All tissues were fixed in 4% phosphate-buffered formaldehyde for a minimum of two weeks before further processing.

### αSyn pathology

αSyn pathology αSyn pathology was assessed using immunohistochemistry (monoclonal antibody 5G4). We examined 12 brain, nine spinal and two peripheral regions (Supplementary Table 1, Supplementary Fig. 4). CNS pathology was scored semi-quantitatively following the third DLBC consensus guidelines,^11^ while peripheral pathology was graded dichotomously due to the generally modest pathology and fewer and inconsistent sampling.^32,36,37^

Subjects were classified into subtypes based on the established staging frameworks using the 0–4 scoring system defined by the third DLBC: The USSLB,^13^ the modified DLBC system by Leverenz and colleagues.^10,11,38^ We opted for the Leverenz modification rather than the original DLBC system due to the high number of unclassifiable cases when applying the latter.

### Alzheimer’s disease and TDP-43 pathology

Alzheimer’s disease neuropathology was evaluated using standard staging systems: A*β* deposition via Thal phase,^39–41^ neuritic plaque density via Consortium to Establish a Registry for Alzheimer’s Disease (CERAD) score,^42,43^ and tau-related NFTs via Braak staging.^30,41^ These measures were integrated into a composite ABC score to classify the severity of Alzheimer’s disease neuropathological change (ADNC) as none, low, intermediate or high.^44^ The likelihood of dementia with Lewy bodies was defined as low, intermediate, or high based on the ABC score and the modified DLBC neuropathology subtypes as outlined by the DLBC consensus guidelines.^10,44^

TAR DNA-binding protein 43 (TDP-43) pathology was staged (1–3) according to limbic-predominant age-related TDP-43 encephalopathy (LATE-NC) criteria, based on involvement of the amygdala, hippocampus and middle frontal gyrus.^45,46^

### SuStaIn: αSyn progression modeling

We employed the Ordinal Subtype and Stage Inference (SuStaIn) to model disease progression.^47,48^ This probabilistic, unsupervised machine-learning algorithm integrates clustering and disease progression modeling to reveal distinct spatial patterns of pathology accumulation on a pseudotemporal axis. SuStaIn models disease progression as a sequential process in which different regions attain specific pathology scores, and it employs Markov Chain Monte Carlo (MCMC) sampling to quantify uncertainty in the inferred sequence.^23,47,49^ Central to this approach is the assumption that pathology follows an ordered process across one or more trajectories. By treating individual cross-sectional postmortem samples as snapshots of different disease stages, SuStaIn reconstructs these continuous progression paths from an assumed baseline to an endpoint.^23,48^

The ordinal version of SuStaIn is designed specifically for ordered rating data, such as neuropathological density scores.^48^ We applied the Ordinal SuStaIn algorithm to αSyn density scores across 19 anatomical regions: Twelve brain, five spinal cord, and two peripheral regions. For each individual, at each spinal level, the dorsal or ventral horn with the greater αSyn pathology was selected for inclusion in the model. Peripheral regions included the adrenal gland and the lumbar DRG. Only individuals with detectable α-synuclein pathology—defined as a pathology density score ≥1 in at least one region—were included in model training (“αSyn+”, *n* = 139). Cases without pathology in any region (“αSyn-”, *n* = 165) were excluded, as they did not contribute information relevant to disease staging.

The model utilized probability estimates for each pathology score, fitted using normal distributions (fixed standard deviation = 0.5) and normalized to sum to one (Supplementary Tables 2–4). ^24,25,50^ Missing density scores were handled using the empirical distribution of the respective region from αSyn+ individuals, minimizing bias compared to case-wise exclusion or conventional imputation methods.^47^ The model underwent training using 100,000 MCMC iterations with 75 start-points, and the evaluation was performed for one to five potential subtypes. As 62 transitions occur across all regions, 62 stages were modeled. Ten-fold cross-validation was performed to calculate the cross-validation information criterion (CVIC) and log-likelihood across folds, with the optimal number of subtypes selected based on the lowest CVIC and highest log-likelihood.^23^ The final model was trained on all αSyn+ individuals (*n* = 139) for the selected number of subtypes. Each individual was assigned a SuStaIn subtype, reflecting their disease progression pattern, and a SuStaIn stage, indicating their position along the pathological trajectory, each with associated confidence measures. For subsequent statistical analyses, only individuals with a SuStaIn stage ≥1 and a high-confidence subtype assignment (probability > 0.5) were included (*n* = 138). Cases with ambiguous subtype classifications due to overlapping progression trajectories (*n* = 15) were excluded from further analysis and designated as “Olf-only” (Supplementary Table 5).

To evaluate model sensitivity, analyses were repeated in individuals with complete data, focusing exclusively on brain and spinal cord regions, while excluding peripheral regions with substantial missing data (50.3% for the adrenal gland and 29.4% for the lumbar dorsal root ganglion among αSyn+ cases). Additionally, the model was rerun using only brain regions. To address high feature dimensionality arising from numerous events, the model was reanalysed with a reduced set of 6 regions (down from 19), decreasing the number of stages from 62 to 21. Results of these sensitivity analyses are presented in Supplementary Figure 3.

### Statistical analyses

Multivariate regression analyses compared αSyn+ (*n* = 139) and αSyn- (*n* = 165) cohorts and evaluated differences across SuStaIn subtypes for demographic, clinical, and pathological variables. Linear regression was applied to continuous outcomes, logistic regression to categorical variables, and ordinal logistic regression to ordinal variables. Models comparing the αSyn+ and αSyn-cohorts were adjusted for age and sex. When evaluating differences among the distinct SuStaIn subtypes, models were further adjusted for SuStaIn stage and corrected for multiple comparisons using the false discovery rate (FDR).^52^ Furthermore, regional αSyn density differences among the three SuStaIn subtypes were systematically assessed across a panel of 19 anatomical regions (comprising 12 brain, 5 spinal cord, and 2 peripheral sites), with these comparisons also undergoing FDR correction for multiple testing.

Global, brain, and spinal αSyn burden were quantified as the summed density scores across the brain and spinal cord, brain only, and spinal cord only, respectively. These metrics were compared across SuStaIn subtypes and correlated with SuStaIn stage using Spearman’s rank correlation and locally estimated scatterplot smoothing (LOESS) regression to assess associations and trends. Univariate Spearman correlations were assessed between SuStaIn stage and existing neuropathological staging systems [USSLB (stages IIa/IIb combined), modified DLBC classifications (brainstem-predominant and amygdala-predominant types grouped as equivalent stages)] or MMSE scores. Model training and all statistical analyses were performed in Python 3.8.10, with two-sided *P* < 0.05 considered nominally significant unless otherwise specified.

## Results

In this population-based autopsy study, we analysed 304 subjects from the Vantaa 85+ cohort. A summary of cohort characteristics is provided in Table 1. 165 individuals exhibited no αSyn pathology in the CNS or periphery (αSyn-), while 139 had at least one region with an αSyn density score ≥ 1 (αSyn+). Compared to αSyn-individuals, αSyn+ subjects had a more severe cognitive impairment (*P* = 0.002) and a higher prevalence of dementia (*P* = 0.004). Additionally, αSyn+ individuals were more likely to exhibit Alzheimer’s co-pathologies and had a higher ABC score (*P* < 0.001). Specifically, they were assigned higher Braak NFT stages (*P* < 0.001), more advanced Thal A*β* plaque phases (*P* < 0.001), and greater CERAD neuritic plaque burden (*P* = 0.011). Additionally, the αSyn+ group showed a higher likelihood of elevated LATE-NC stage (*P* = 0.005). Although the αSyn+ group had a higher proportion of APOE ε4 allele carriers, this difference was not significant (*P* = 0.054). No differences were observed between the two groups in terms of sex distribution (*P* = 0.457), age at death (*P* = 0.652), or dementia duration (*P* = 0.492).

**Table 1.**
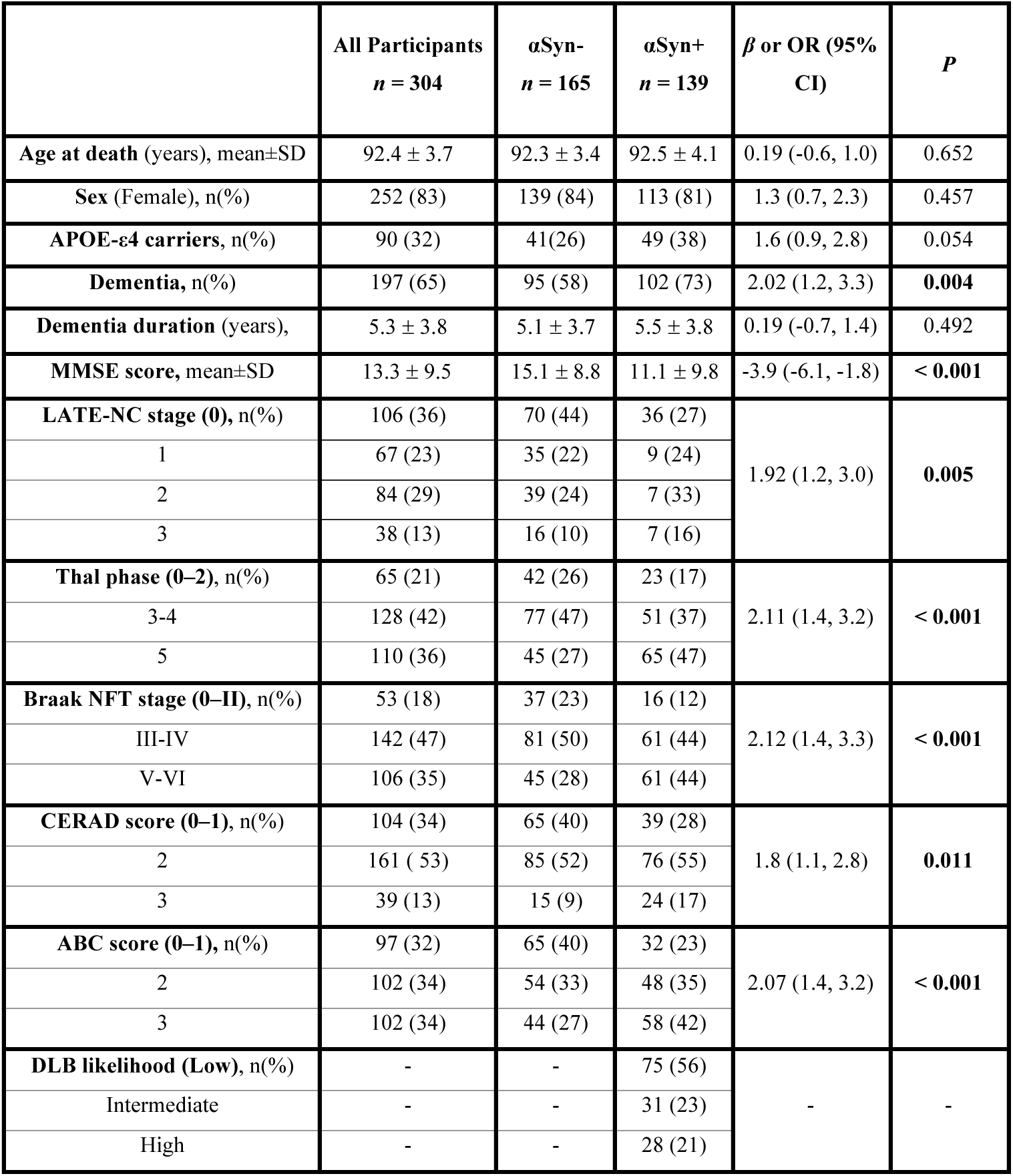
Demographic, clinical, and neuropathological characteristics of the Vantaa 85+ cohort. Ordinal logistic regression was applied to ordered categorical variables, logistic regression to binary variables, and linear regression to continuous variables. Results of subtype comparisons are reported as odds ratios (OR, 95% CI) for ordinal and logistic models, and beta coefficients (*β*, 95% CI) for linear models. αSyn, α-synuclein; CERAD, Consortium to Establish a Registry for Alzheimer’s Disease; MMSE, Mini-Mental State Examination; LATE-NC, limbic-predominant age-related TDP-43 encephalopathy; NFT, neurofibrillary tangle.

### Data-driven subtyping and staging of αSyn propagation

Using Ordinal SuStaIn, we modeled the spatiotemporal progression of αSyn pathology based on density scores from 12 brain, nine spinal cord, and two peripheral regions in αSyn+ individuals. Model-fit statistics, including log-likelihood and 10-fold cross-validation, indicated that three distinct spatiotemporal subtypes best described the data (Supplementary Fig. 1). We termed these subtypes Olfactory-Amygdala (“Olf-Amy”), Olfactory-Brainstem (“Olf-Brainstem”), and Dorsal Motor Nucleus of Vagus-Thoracic Intermediolateral Column (“DMV-TIML”) (Fig. 1). They each exhibited unique sequences of αSyn accumulation across the assessed regions. Reflecting the population-based nature of the cohort, stage distribution skewed toward earlier stages, with relatively even distribution of mid-stage cases, and sparse late-stage cases (Supplementary Fig. 2). Consequently, early-stage pathology trajectories were well-resolved, whereas late-stage trajectories were less precisely defined due to limited data, as reflected by the more saturated colors in early stages and less saturated colors in late stages in positional variance diagrams (Fig. 1B).

**Figure 1.**
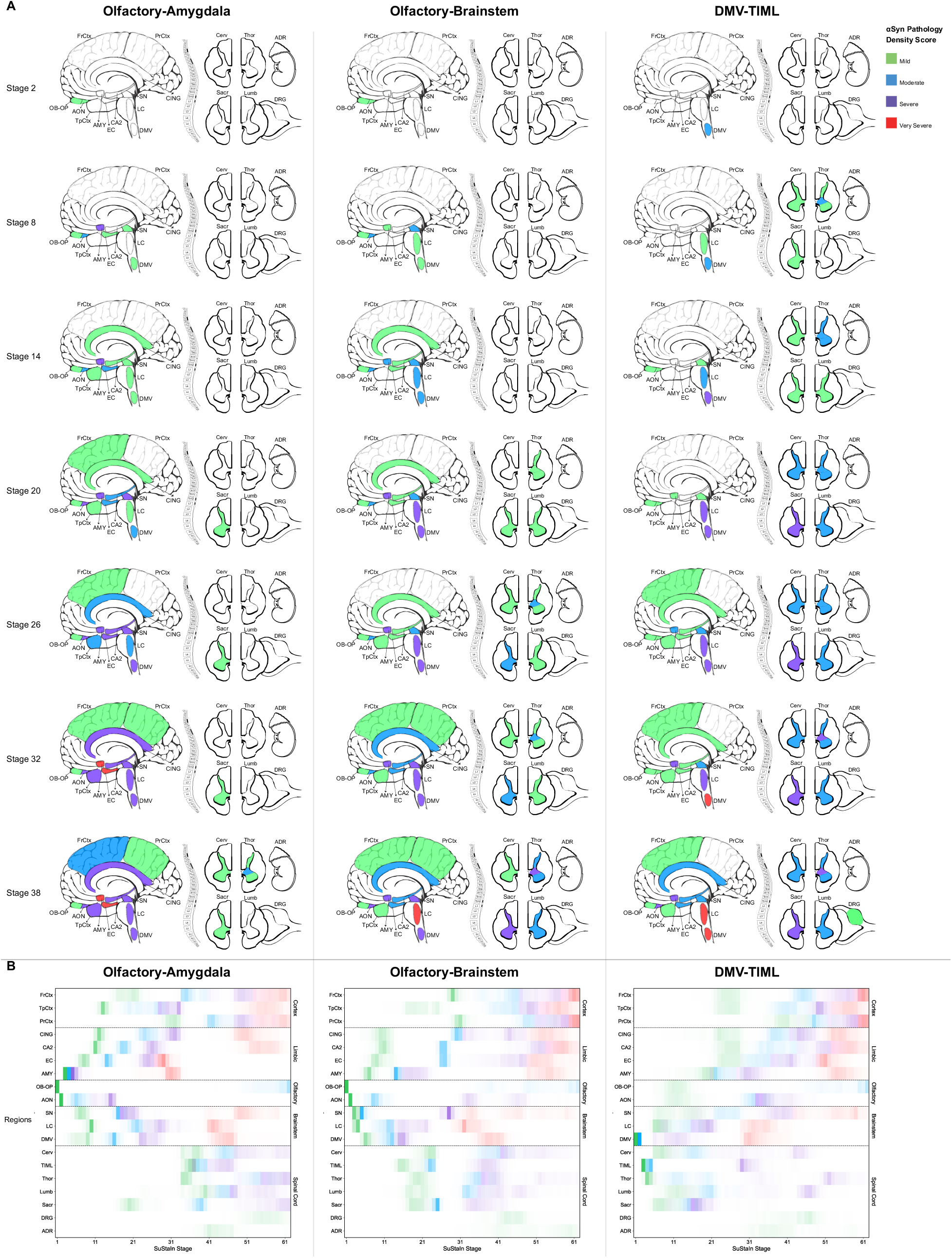
SuStaIn-inferred subtypes of αSyn progression. The SuStaIn algorithm identified three distinct trajectories of αSyn progression: Olf-Amy (left), Olf-Brainstem (middle), and DMV-TIML (right). (**A**) αSyn deposition maps are shown for SuStaIn stages 2 to 38 at 6-stage intervals. Certain anatomical regions are displayed larger than their actual size for demonstration purposes to clearly show the severity mapping. (**B**) Each box in the positional variance diagrams depicts the certainty of regional involvement, with darker shading indicating higher certainty. In both panels, color hue represents the severity of αSyn pathology: green = mild, blue = moderate, purple = severe, and red = very severe; while white boxes indicate no transition in severity. ADR, adrenal gland; AON, anterior olfactory nucleus; AMY, amygdala; CA2, cornu ammonis 2; Cerv, cervical spinal cord; CING, cingulate gyrus; DMV, dorsal motor nucleus of the vagus; DRG, dorsal root ganglion; EC, entorhinal cortex; FrCtx, frontal cortex; LC, locus coeruleus; Lumb, lumbar spinal cord; OB-OP, peripheral olfactory bulb/peduncle; Olf, olfactory; PrCtx, parietal cortex; SN, substantia nigra; Sacr, sacral spinal cord; Thor, thoracic spinal cord; TIML, thoracic intermediolateral column; TpCtx, temporal cortex; SuStaIn, Subtype and Stage Inference.

Each individual was assigned a SuStaIn subtype and stage (0-62) accompanied by an uncertainty estimate. Of the 139 αSyn+ individuals, one was assigned a SuStaIn stage of 0 with a subtype probability below 50%, indicating insufficient pathology for confident subtyping, and was excluded from further analysis. Initial subtype classification identified 60 individuals as Olf-Amy, 41 as Olf-Brainstem, and 37 as DMV-TIML. However, detailed examination revealed overlapping early pathology patterns between Olf-Amy and Olf-Brainstem, both originating in the OB-OP at stage 1 and progressing to the AON at stage 2 (Fig. 1). Consequently, 15 individuals with pathology confined to olfactory regions were initially assigned to the more prevalent Olf-Amy subtype but exhibited low assignment probabilities (50.5%–51.6%, Supplementary Fig. 2). To mitigate potential bias in downstream analyses, these cases were reclassified as an independent “Olf-only” group and excluded from further statistical comparisons, with their demographic, clinical, and neuropathological information provided in Supplementary Table 5 and Supplementary Figure 4. Following this adjustment, the final subtype distribution comprised 45 (32.6%) individuals in Olf-Amy, 41 (29.7%) in Olf-Brainstem, 37 (26.8%) in DMV-TIML, and 15 (10.8%) in Olf-only. Altogether, SuStaIn analysis suggested that 73.2% of individuals had an olfactory origin, while 26.8% originated in the DMV. Subtype assignment probabilities for these individuals were robust and consistent across early, mid, and late stages, supporting the reliability of the subtype classifications (Supplementary Fig. 2).

### Divergent origin and spread of αSyn pathology identified by the SuStaIn model

The SuStaIn model identified three distinct spatiotemporal progression patterns of αSyn pathology, as illustrated in Figure 1. The Olf-Amy and Olf-Brainstem subtypes have an initial site of αSyn pathology in the OB-OP at stage 1, progressing to the AON at stage 2, with AON pathology reaching moderate severity by stages 6-8. Mild and moderate DMV pathology emerged later in these subtypes, at stages 6–9 and 13–16, respectively. Conversely, the DMV-TIML subtype exhibited initial pathology in the DMV at stage 1 (mild) and stage 2 (moderate), with olfactory involvement delayed until stage 11 and AON reaching moderate severity by stage 32.

Despite a shared olfactory origin, Olf-Amy and Olf-Brainstem trajectories diverged. The Olf-Amy subtype progressed from the olfactory regions to the amygdala, followed by the brainstem and limbic regions, before spreading to the neocortex and, lastly, the spinal cord. Conversely, the Olf-Brainstem subtype advanced from the olfactory regions to the brainstem, then to the amygdala and limbic areas, with spinal cord involvement preceding neocortical spread in the final stages. The DMV-TIML subtype followed a distinct caudorostral progression pattern. Originating in the DMV, followed by the TIML and spinal cord horns, the pathology ascended sequentially to the higher brainstem, then to the olfactory regions, amygdala, limbic structures, and, ultimately, the neocortex. Peripheral regions, including the adrenal gland and lumbar DRG, were the last to exhibit αSyn pathology across all subtypes, with the DMV-TIML subtype showing slightly earlier adrenal involvement compared to the Olf-Amy and Olf-Brainstem subtypes.

Figure 2 presents the spatial distribution of αSyn pathology scores across the brain, spinal cord, and peripheral regions for each SuStaIn subtype, shown both overall (Fig. 2A) and stratified by stage (Fig. 2B). In the Olf-Amy subtype, αSyn pathology was consistently observed in olfactory and limbic regions, with all 45 individuals showing involvement of the OB-OP, AON, and amygdala. Notably, only one individual exhibited mild pathology in the amygdala, and four showed mild involvement of the AON. In the Olf-Brainstem subtype, all 41 individuals displayed pathology in the OB-OP and substantia nigra, with locus coeruleus and AON affected in all but one case. Among the 37 individuals classified as DMV-TIML, pathology was present in the DMV in all cases, with 31 also showing involvement of the TIML. Peripheral pathology was most prominent in the DMV-TIML subtype, with adrenal and DRG involvement observed in 56.5% and 43.6% of individuals with complete data, respectively, compared to 27.8% and 22.6% in Olf-Brainstem, and 20.0% and 6.0% in Olf-Amy.

**Figure 2.**
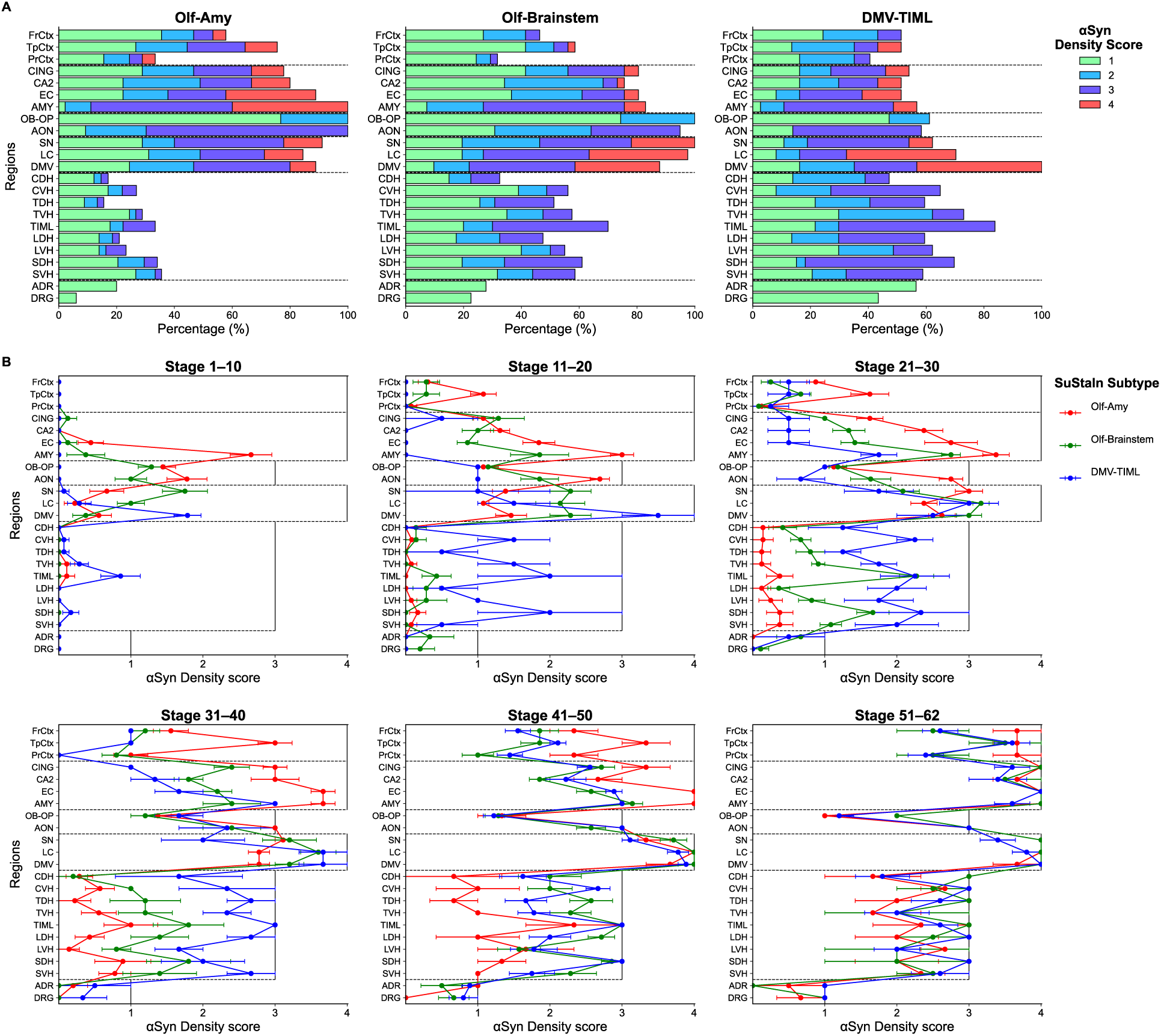
Spatial distribution of αSyn pathology across SuStaIn subtypes. **(A)** Percentage stacked bar charts showing the distribution of αSyn pathology density scores across brain, spinal cord, and peripheral regions in individuals assigned to Olf-Amy (left), Olf-Brainstem (Middle), and DMV-TIML (right) subtypes. **(B)** Mean αSyn density scores with standard deviation error bars across the six SuStaIn stage groups (SuStaIn stages 1–10, 11–20, …, 51–62) for each subtype: Olf-Amy (red), Olf-Brainstem (green), and DMV-TIML (blue). ADR, adrenal gland; AON, anterior olfactory nucleus; AMY, amygdala; CA2, cornu ammonis 2; Cerv, cervical spinal cord; CING, cingulate gyrus; DMV, dorsal motor nucleus of the vagus; DRG, dorsal root ganglion; EC, entorhinal cortex; FrCtx, frontal cortex; LC, locus coeruleus; Lumb, lumbar spinal cord; OB-OP, peripheral olfactory bulb/peduncle; Olf, olfactory; PrCtx, parietal cortex; SN, substantia nigra; Sacr, sacral spinal cord; Thor, thoracic spinal cord; TIML, thoracic intermediolateral column; TpCtx, temporal cortex; SuStaIn, Subtype and Stage Inference.

SuStaIn stage assignments were comparable across all subtypes (all pairwise comparisons *P* > 0.05; Fig. 3A) and demonstrated near-perfect collinearity with global αSyn burden (*r* = 0.98; Fig. 3D). Despite this global similarity, regional accumulation patterns varied. In the brain, αSyn burden was highest in the Olf-Amy subtype, followed by Olf-Brainstem and DMV-TIML (Fig. 3B). While the Olf-Amy subtype appeared to exhibit an early trajectory of burden accumulation (Fig. 3E), these trends did not reach statistical significance. Conversely, spinal cord pathology showed significant divergence: spinal αSyn burden was markedly higher in both the Olf-Brainstem and DMV-TIML subtypes compared to Olf-Amy (Table 2; Fig. 3C). Specifically, the DMV-TIML subtype was characterized by a rapid rise to an early plateau, contrasting with the steady increase in the Olf-Brainstem group and the delayed onset observed in the Olf-Amy group (Fig. 3F).

**Figure 3.**
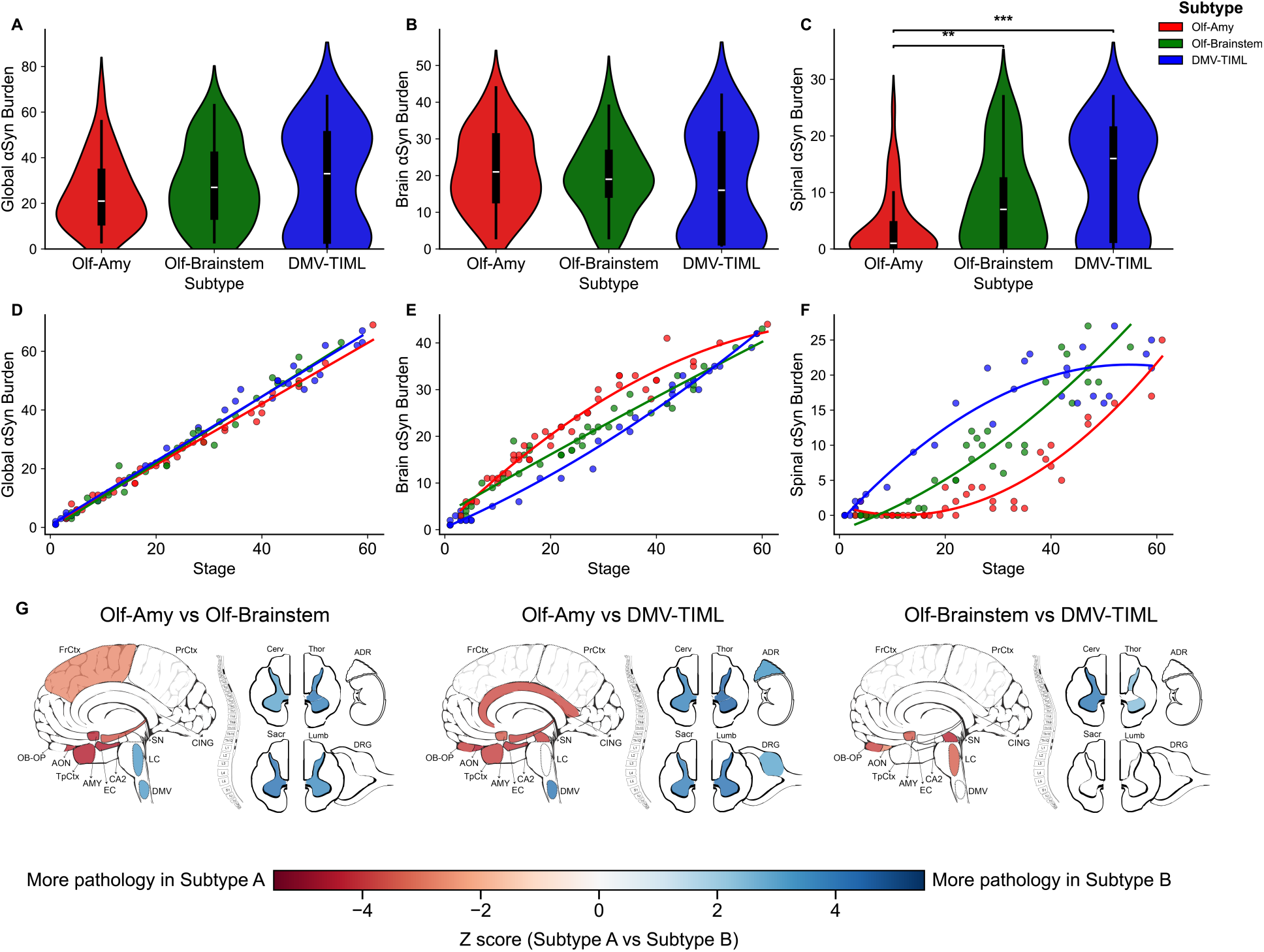
Comparison of αSyn burden across SuStaIn subtypes. (**A–C**) Violin plots showing global (left), brain (middle), and spinal (right) αSyn burden in Olf-Amy (red), Olf-Brainstem (green), and DMV-TIML (blue) subtypes. (**D–F**) LOESS regression scatterplots depicting the relationship between SuStaIn stage and global (left), brain (middle), and spinal (right) αSyn burden for Olf-Amy (red), Olf-Brainstem (green), and DMV-TIML (blue) subtypes. (**G**) Regional maps of pairwise ordinal logistic regression results, corrected for multiple comparisons, for Olf-Amy vs Olf-Brainstem (left), Olf-Amy vs DMV-TIML (middle), and Olf-Brainstem vs DMV-TIML (right). Colors indicate z-scores from regression analyses, with red representing higher pathology in subtype A and blue representing higher pathology in subtype B (for A vs B comparisons); while white indicates no significant difference. For demonstrative clarity of the severity mapping, certain anatomical areas have been enlarged past their actual size. Boxplots display the median and interquartile range, with whiskers extending to the minimum and maximum values. αSyn, α-synuclein; DMV-TIML, dorsal motor nucleus of the vagus–thoracic intermediolateral nucleus; LOESS, locally estimated scatterplot smoothing; Olf-Amy, olfactory-amygdala; Olf-Brainstem, olfactory-brainstem; SuStaIn, Subtype and Stage Inference. Statistical significance is indicated as ∗p<0.05, ∗∗p<0.01, and ∗∗∗p<0.001 (FDR-corrected).

**Table 2.** Characteristics of SuStaIn-derived αSyn subtypes. Ordinal logistic regression was applied to ordered categorical variables, logistic regression to binary variables, and linear regression to continuous variables. Results of subtype comparisons are reported as odds ratios (OR, 95% CI) for ordinal and logistic models, and beta coefficients (*β*, 95% CI) for linear models. Statistical significance is indicated as ∗p<0.05, ∗∗p<0.01, and ∗∗∗p<0.001 (FDR-corrected). αSyn, α-synuclein; CERAD, Consortium to Establish a Registry for Alzheimer’s Disease; DLB, dementia with Lewy bodies; DMV-TIML, dorsal motor nucleus of the vagus–thoracic intermediolateral nucleus; LATE-NC, limbic-predominant age-related TDP-43 encephalopathy; MMSE, Mini-Mental State Examination; NFT, neurofibrillary tangle; Olf-Amy, olfactory-amygdala; Olf-Brainstem, olfactory-brainstem; SD, standard deviation; SuStaIn, Subtype and Stage Inference.

|  | SuStaIn Subtype |  |  | SuStaIn Subtype Comparison |  |  |
| --- | --- | --- | --- | --- | --- | --- |
| | Olf-Amy<br>(n=45) | Olf-Brainstem<br>(n=41) | DMV-TIML<br>(n=37) | Olf-Brainstem vs<br>Olf-Amy, $\beta$ or OR<br>(95% CI) | DMV-TIML vs<br>Olf-Amy, $\beta$ or OR<br>(95% CI) | DMV-TIML vs<br>Olf-Brainstem, $\beta$<br>or OR (95% CI) |
| Age at death (years),<br>mean±SD | 92.7 ± 4.0 | 91.5 ± 3.4 | 93.1 ± 4.7 | 1.1 (-0.6, 2.9) | -0.4 (-2.1, 1.4) | -1.5 (-3.3, 0.3) |
| Sex (Female), n(%) | 38 (84) | 30 (73) | 30 (81) | 2.0 (0.7, 5.7) | 1.2 (0.4, 4.0) | 0.6 (0.2, 1.9) |
| APOE-ε4 carriers, n(%) | 24 (59) | 11 (31) | 10 (28) | 3.8 (1.4, 10.2)* | 3.7 (1.4, 9.8)* | 1.0 (0.3, 2.8) |
| Dementia, n(%) | 41 (91) | 26 (63) | 23 (62) | 6.0 (1.7, 21.6)* | 7.7 (2.1, 28.7)** | 1.3 (0.4, 3.6) |
| Dementia duration<br>(years), mean±SD | 6.8 ± 3.8 | 3.1 ± 2.1 | 5.2 ± 3.7 | 3.5 (1.7, 5.2)*** | 1.8 (0.1, 3.6) | -1.6 (-3.6, 0.3) |
| Brain αSyn burden,<br>mean±SD | 21.6 ± 10.7 | 19.8 ± 9.9 | 17.4 ± 14.7 | 3.7 (2.7, 4.8)*** | 6.1 (5.0, 7.1)*** | 2.3 (1.2, 3.4)*** |
| Spinal αSyn burden,<br>mean±SD | 3.8 ± 5.9 | 8.5 ± 8.5 | 12.2 ± 9.5 | -4.6 (-6.2, -3.0)*** | -7.6 (-9.2, -5.9)*** | -3.0 (-4.7, -1.3)*** |
| MMSE score, mean±SD | 6.4 ± 8.9 | 15.7 ± 8.6 | 13.1 ± 9.9 | -7.8 (-11.7, -3.9)*** | -6.7 (-10.6, -2.9)** | 1.1 (-2.9, 5.1) |
| LATE-NC stage (0), n(%) | 7 (16) | 15 (39) | 12 (34) | 2.31 (1.03, 5.18) | 1.97 (0.87, 4.46) | 0.85 (0.36, 2.03) |
| 1 | 11 (24) | 10 (26) | 9 (26) |  |  |  |
| 2 | 20 (44) | 10 (26) | 7 (20) |  |  |  |
| 3 | 7 (16) | 4 (10) | 7 (20) |  |  |  |
| Thal phase (0–2), n(%) | 0 (0) | 14 (34) | 6 (16) | 16.5 (6.0, 45.5)*** | 12.0 (4.5, 32.1)*** | 0.7 (0.3, 1.7) |
| 3–4 | 9 (20) | 15 (37) | 23 (62) |  |  |  |
| 5 | 36 (80) | 12 (29) | 8 (22) |  |  |  |
| Braak NFT stage (0–II),<br>n(%) | 0 (0.0) | 9 (23) | 6 (16) | 26.8 (9.0, 80.0)*** | 18.5 (6.4, 53.1)*** | 0.7 (0.3, 1.7) |
| III–IV | 9 (20) | 24 (60) | 24 (65) |  |  |  |
| V–VI | 36 (80) | 7 (18) | 7 (19) |  |  |  |
| CERAD score (0–1), n(%) | 1 (2) | 21 (51) | 12 (32) | 30.2 (8.7, 105.1)*** | 17.2 (5.2, 57.3)*** | 0.6 (0.2, 1.4) |
| 2 | 26 (58) | 17 (42) | 25 (68) |  |  |  |
| 3 | 18 (40) | 3 (7) | 0 (0) |  |  |  |
| ABC score (0–1), n(%) | 0 (0) | 19 (48) | 9 (24) | 43.8 (14.2, 134.9)*** | 17.3 (6.2, 48.5)*** | 0.4 (0.2, 1.0) |
| 2 | 9 (20) | 15 (37) | 21 (57) |  |  |  |
| 3 | 36 (80) | 6 (15) | 7 (19) |  |  |  |
| DLB likelihood (Low),<br>n(%) | 27 (63) | 13 (33) | 20 (54) | 0.1 (0.0, 0.3)*** | 0.5 (0.2, 1.4) | 4.7 (1.5, 14.9)* |
| Intermediate | 15 (35) | 10 (25) | 6 (16) |  |  |  |
| High | 1 (2) | 16 (41) | 11 (29) |  |  |  |

We also performed pairwise statistical comparisons for regional αSyn density scores across the three SuStaIn subtypes, controlling for stage and correcting for multiple comparisons (Fig. 3G). The Olf-Amy subtype showed significantly higher αSyn burden in limbic, olfactory, and neocortical regions. In contrast, the Olf-Brainstem and DMV-TIML subtypes exhibited greater pathology in the lower brainstem and spinal cord. The DMV-TIML subtype also demonstrated a significantly higher burden of peripheral αSyn pathology compared to Olf-Amy. Between Olf-Brainstem and DMV-TIML, the former showed greater pathology in olfactory structures, the amygdala, and the upper brainstem, whereas the latter was characterized by more extensive αSyn involvement in the upper spinal cord. No significant differences in DMV pathology were observed between these two subtypes. Among peripheral regions, the lumbar DRG was the only area to show a difference, with higher involvement in DMV-TIML compared to Olf-Amy.

### SuStaIn-derived trajectories align with established αSyn staging systems

αSyn+ individuals were classified using two established neuropathological staging frameworks informed by the third DLB guidelines: The USSLB ^13^ and the modified DLBC system by Leverenz ^10,38^. All individuals were assigned to a distinct stage or type under these systems, except for three cases deemed unclassifiable according to the modified DLBC criteria. There was a monotonic association between ordinal stages from both classifications and SuStaIn-derived progression stages (USSLB: *r* = 0.87, *P* < 0.001; modified DLBC: *r* = 0.90, *P* < 0.001; Fig. 4). Early USSLB stages (I, IIa, IIb) and modified DLBC olfactory-only, amygdala-predominant, and brainstem-predominant types mapped predominantly to earlier SuStaIn stages, while mid-stage USSLB III and modified DLBC limbic cases aligned with mid-range SuStaIn stages. Late-stage USSLB IV and modified DLBC neocortical cases corresponded to the latest SuStaIn stages. To assess whether SuStaIn subtypes captured anatomical distinctions beyond stage alone, we examined subtype assignments among individuals in early SuStaIn stages (< 31). Among USSLB stage IIa (brainstem-predominant; *n* = 32) and modified DLBC brainstem-predominant cases (*n* = 30), the majority were assigned to either the DMV-TIML (19 and 18, respectively) or Olf-Brainstem (12 and 10) SuStaIn subtypes. In contrast, all USSLB stage IIb (limbic-predominant; *n* = 14) and modified DLBC amygdala-predominant (*n* = 6) individuals were assigned to the Olf-Amy subtype. Individuals in USSLB stage III and modified DLBC limbic stages were distributed across Olf-Brainstem (15 and 16, respectively) and Olf-Amy (8 and 20), whereas all USSLB stage IV cases (*n* = 7) were classified exclusively as subtype Olf-Amy. In summary, the data-driven SuStaIn classification aligned closely with two commonly used staging schemes for synucleinopathy.

**Figure 4.**
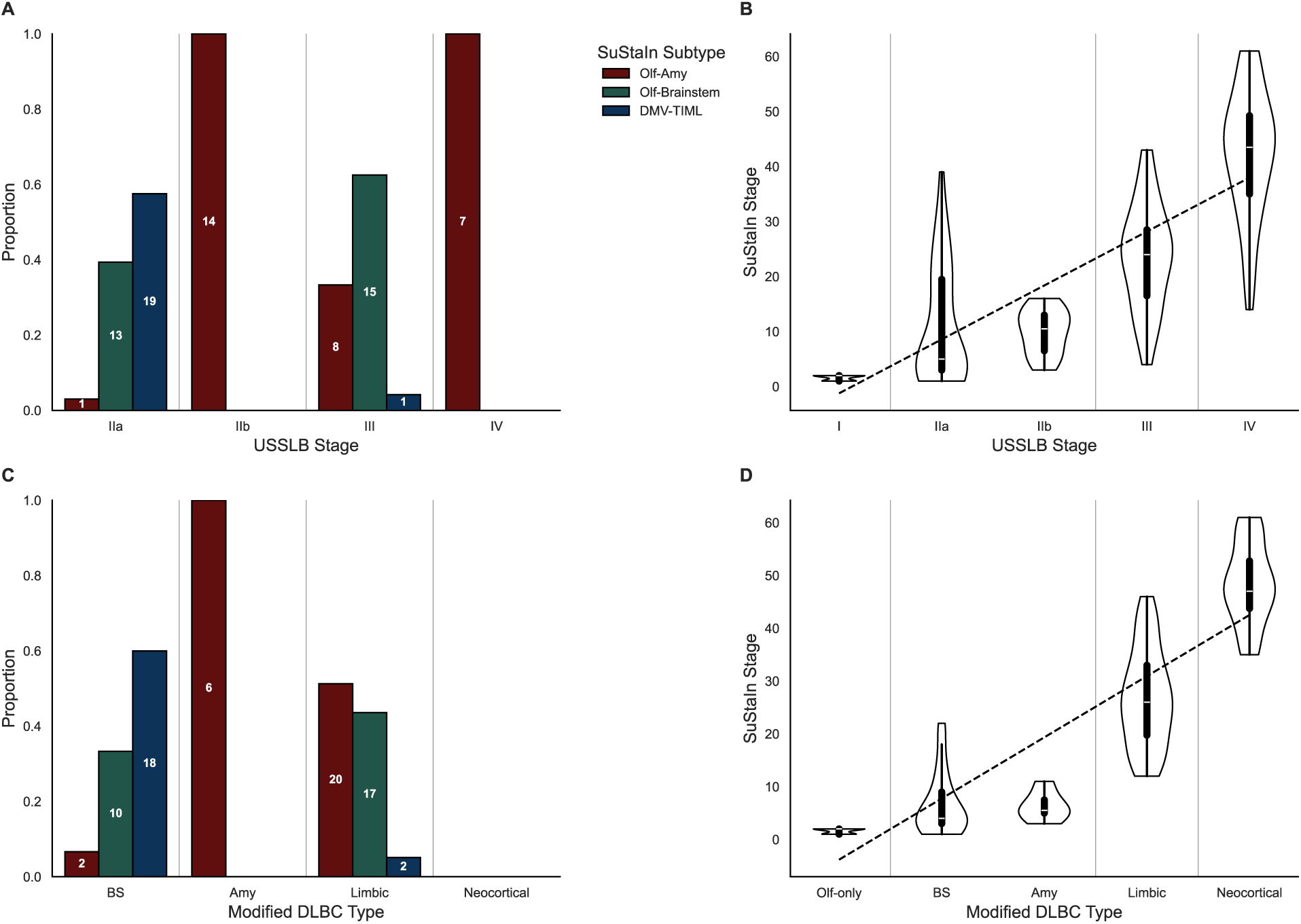
Comparison of SuStaIn-inferred subtypes with established staging systems. The proportion of early-stage individuals (SuStaIn stage <31) assigned to USSLB **(A)** and modified DLBC **(C)** stages across Olf-Amy (red), Olf-Brainstem (green), and DMV-TIML (blue) subtypes. Spearman correlations are shown between SuStaIn stage and USSLB stage (**B**; IIa and IIb combined) and between SuStaIn stage and modified DLBC type (**D**; brainstem-predominant and amygdala-predominant combined). Boxplots display the median and interquartile range, with whiskers extending to the minimum and maximum values. AMY, amygdala-predominant; BS, brainstem-predominant; DLBC, Dementia with Lewy Bodies Consortium; DMV-TIML, dorsal motor nucleus of the vagus–thoracic intermediolateral nucleus; Olf-Amy, olfactory-amygdala; Olf-Brainstem, olfactory-brainstem; Olf-only, olfactory bulb-only; USSLB, Unified Staging System for Lewy Body Disorders.

### SuStaIn subtypes reveal distinct genetic, neuropathological, and clinical profiles

We next interrogated demographic, genetic, and neuropathological features across SuStaIn-defined αSyn subtypes (Fig. 5, Table 2). No significant differences in sex distribution or age at death were observed between subtypes. Furthermore, age at death was not associated with SuStaIn stage (*r* = −0.05, *P* = 0.596), brain αSyn burden (*r* = −0.09, *P* = 0.310), or spinal cord αSyn burden (*r* = −0.02, *P* = 0.794), likely reflecting the age-constrained nature of the cohort. These associations were not observed within each subtype (Olf-Amy: *r* = −0.22 to −0.13; Olf-Brainstem: *r* = −0.11 to 0.01; DMV-TIML: *r* = 0.11 to 0.18; all *P* > 0.05).

**Figure 5.**
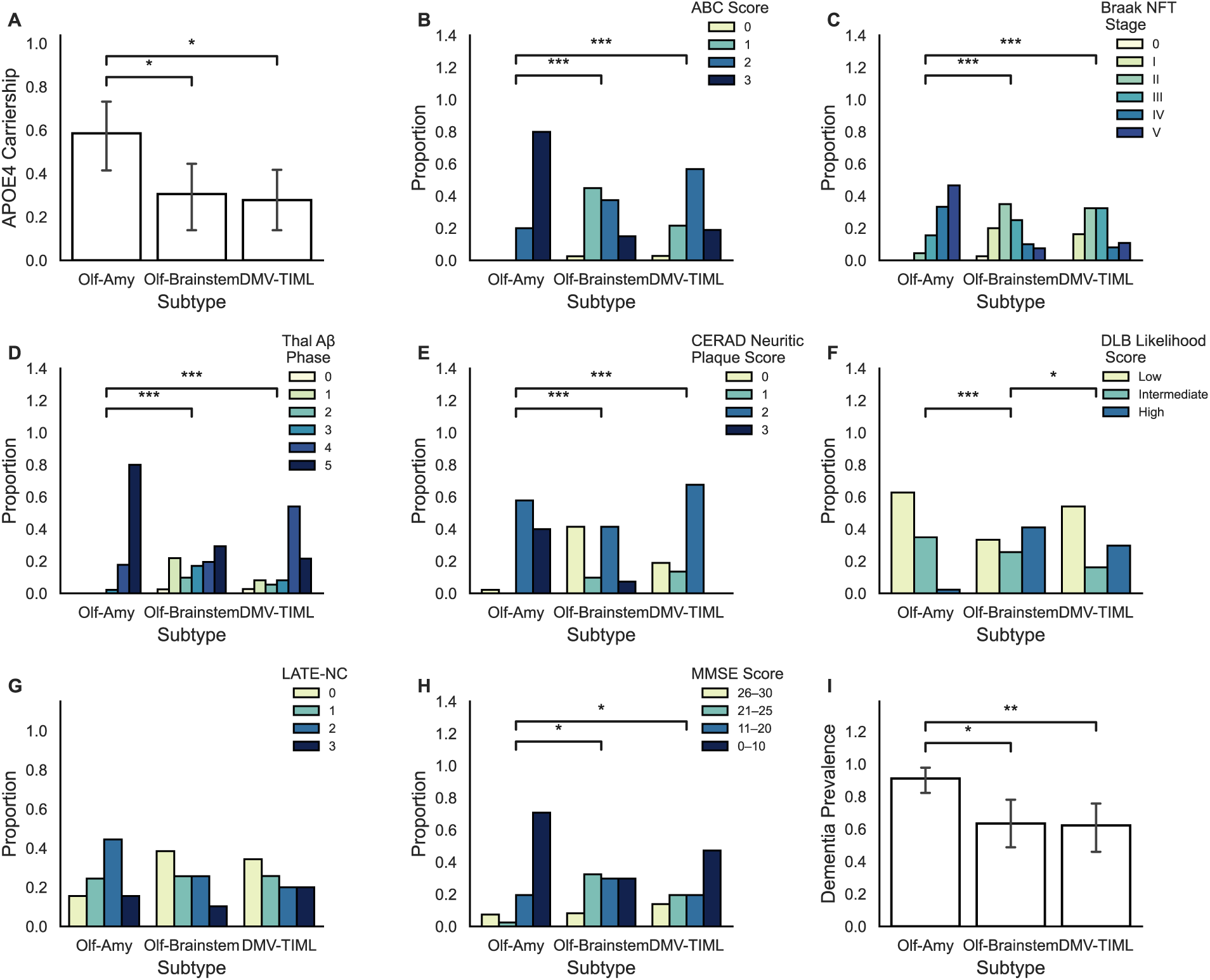
Comparisons of SuStaIn subtypes across clinical, neuropathological, and genetic metrics. (**A**) APOE carrier status, (**B**) ABC score, (**C**) Braak NFT stage, (**D**) Thal A*β* phase, (**E**) CERAD neuritic plaque score, (**F**) DLB likelihood score, (**G**) LATE-NC stage, (**H**) MMSE score, and (**I**) dementia prevalence. A*β*, amyloid-*β*; APOE, apolipoprotein E; CERAD, Consortium to Establish a Registry for Alzheimer’s Disease; DLB, dementia with Lewy bodies; DMV-TIML, dorsal motor nucleus of the vagus–thoracic intermediolateral nucleus; LATE-NC, limbic-predominant age-related TDP-43 encephalopathy; MMSE, Mini-Mental State Examination; NFT, neurofibrillary tangle; Olf-Amy, olfactory-amygdala; Olf-Brainstem, olfactory-brainstem; SuStaIn, Subtype and Stage Inference. Statistical significance is indicated as ∗p<0.05, ∗∗p<0.01, and ∗∗∗p<0.001 (FDR-corrected).

Genotypic analysis revealed differences in APOE ε4 carriership across subtypes. Olf-Amy individuals exhibited a significantly higher frequency of APOE ε4 alleles (59%) compared to Olf-Brainstem (31%) and DMV-TIML (28%) subtypes (Fig. 5A). Neuropathological assessments revealed a significantly greater burden of Alzheimer’s co-pathology among Olf-Amy cases. Olf-Amy cases exhibited a higher ADNC as indicated by the ABC score compared to the other subtypes (all OR > 17.0, *P* < 0.001). Conversely, no significant differences in ADNC were observed between the Olf-Brainstem and DMV-TIML groups. All Olf-Amy cases met criteria for intermediate (20%) or high (80%) ADNC. In contrast, Olf-Brainstem individuals were predominantly intermediate (46%) or low (37%), and DMV-TIML cases were largely low (57%) or intermediate (24%) (Fig. 5B). With respect to tau pathology, Olf-Amy individuals were significantly more likely to fall within advanced Braak stages (III–IV: 20%; V–VI: 80%) compared to Olf-Brainstem (Braak III–IV: 59%) and DMV-TIML (Braak III–IV: 65%) (Fig. 5C). Similarly, Thal A*β* staging was significantly elevated in Olf-Amy cases, with all individuals in phase 3–4 (20%) or phase 5 (80%). In contrast, the DMV-TIML group was primarily concentrated in phases 3–4 (62%), while Olf-Brainstem individuals showed a broader distribution across Thal phases (Fig. 5D). CERAD scoring further supported the greater A*β* burden in Olf-Amy, with nearly all cases exhibiting moderate (58%) or frequent (40%) neuritic plaques. This was significantly greater than in Olf-Brainstem and DMV-TIML, which showed more variable plaque density (no/sparse: 51% and 32%, moderate: 41% and 67%, respectively) (Fig. 5E).

While Olf-Amy cases displayed neuropathological profiles consistent with concomitant Alzheimer’s disease, Olf-Brainstem individuals were more likely to meet neuropathological criteria for dementia with Lewy bodies.^10^ Specifically, Olf-Brainstem subjects exhibited higher DLB likelihood scores compared to both Olf-Amy and DMV-TIML subtypes, with 63% of the cohort categorized as having intermediate or high likelihood. In contrast, the majority of Olf-Amy (60%) and DMV-TIML (54%) cases were classified as low DLB likelihood (Fig. 5F). Assessment of TDP-43 pathology revealed no significant differences across subtypes (*P* > 0.05). Although a higher proportion of Olf-Amy cases exhibited stage 2 pathology (44%) compared to Olf-Brainstem (26%) and DMV-TIML (20%), pairwise comparisons did not reach statistical significance (Fig. 5G).

The divergent neuropathological patterns across subtypes were mirrored by distinct clinical profiles. The Olf-Amy group exhibited the most profound cognitive impairment, with 80% of individuals categorized as having moderate-to-severe impairment based on MMSE scores. This proportion was significantly higher than that observed in the Olf-Brainstem (55%) and DMV-TIML (64%) subtypes (Fig. 5H). Consistent with these cognitive deficits, dementia prevalence was highest in the Olf-Amy group (91%), significantly exceeding the rates in both Olf-Brainstem (63%) and DMV-TIML (62%) cases (Fig. 5I). Furthermore, the Olf-Amy subtype was characterized by a significantly longer mean dementia duration (6.8 years) compared to the Olf-Brainstem group (3.1 years). However, dementia duration did not differ significantly between the Olf-Amy and DMV-TIML subtypes (5.2 years).

### Cognitive performance reflects the spatiotemporal distribution of αSyn and Alzheimer’s disease co-pathology

To delineate the independent contributions of αSyn and Alzheimer’s disease pathology to cognitive impairment, we assessed whether cognitive status was exclusively attributable to co-existing ADNC or whether αSyn burden, as indexed by SuStaIn stage, exerted additional effects. We stratified individuals by ADNC severity as indicated by ABC score (low/none, intermediate, high), LATE–NC stage and αSyn status [αSyn−, early-stage αSyn+ (SuStaIn Stage <31), late-stage αSyn+ (SuStaIn Stage ≥ 31)] (Fig. 6). In the regression analyses, however, αSyn burden was modeled continuously using SuStaIn stage. Dementia likelihood was associated with ADNC level, LATE-NC stage and αSyn burden. Crucially, the effects of these three variables were independent and non-interactive, indicating that αSyn burden (SuStaIn stage) remained an independent predictor of dementia [OR = 1.03, 95% CI (1.01, 1.05), *P* = 0.006], after accounting for co-pathology (Supplementary Table 6).

**Figure 6.**
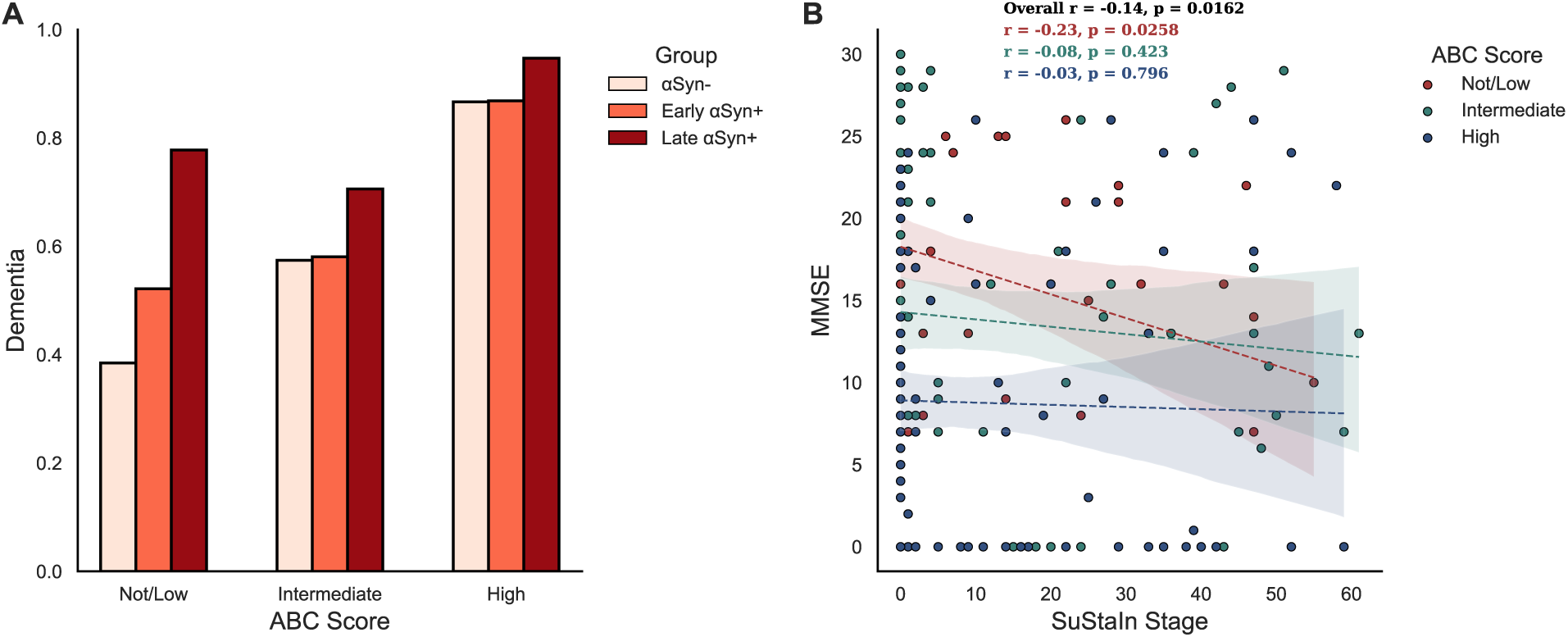
Impact of αSyn and Alzheimer’s disease co-pathology on cognitive outcomes. (**A**) Dementia prevalence across ABC score categories (not/low, intermediate, high) stratified by synucleinopathy status: αSyn-, early αSyn+ (SuStaIn stage <31), and late αSyn+ (SuStaIn stage ≥31). (**B**) Scatterplots showing MMSE scores as a function of SuStaIn stage, with fitted regression lines and 95% confidence intervals for each ABC score category. αSyn, α-synuclein; MMSE, Mini-Mental State Examination; SuStaIn, Subtype and Stage Inference.

Among αSyn+ individuals with no or low ABC pathology (*n* = 28), 16 were demented. These subjects had higher SuStaIn stages and greater corticolimbic αSyn burden compared with non-demented counterparts (Supplementary Fig. 5). When moving from None/Low to Intermediate ADNC, the αSyn− group showed a larger increase in dementia prevalence (38.5% → 57.5%), the early-stage αSyn+ group exhibited a modest rise (52.1% → 58.1%), and the late-stage αSyn+ group showed a slight decrease (77.8% → 70.6%). Progression from intermediate to high ADNC was associated with a greater increase in dementia prevalence across all αSyn groups: αSyn− (57.4% → 86.7%), early-stage αSyn+ (58.1% → 86.8%), and late-stage αSyn+ (70.6% → 94.7%) (Fig. 6A). MMSE scores were similarly influenced by ADNC, LATE–NC stage and αSyn burden (SuStaIn stage: *β* = –0.06, 95% CI (–0.12, –0.01), *P* = 0.048). While no interaction between ADNC and αSyn was observed, the negative correlation between αSyn burden and MMSE was more pronounced in the none/low ADNC group (*r* = −0.23) than in the intermediate (*r* = −-0.08) or high (*r* = −0.03) groups (Fig. 6B). Sensitivity analyses using corticolimbic αSyn burden in place of SuStaIn stage recapitulated these results (Supplementary Table 6). Collectively, these findings indicate that αSyn, Alzheimer’s and TDP-43 pathologies independently contribute to cognitive impairment with an additive effect.

## Discussion

In this population-based autopsy study of 304 individuals aged 85 years and older, we focused on 139 individuals exhibiting αSyn pathology and employed data-driven disease progression modeling to delineate the spatiotemporal progression trajectories of αSyn pathology. We identified three distinct αSyn progression subtypes, each exhibiting unique regional sequences of pathological accumulation. Our SuStaIn analysis suggests that the majority of individuals (73.2%) exhibited initial αSyn deposition in olfactory regions, followed by progression to either the amygdala (32.6%) or brainstem (29.7%). A third subset (26.8%) demonstrated pathology initiation in the DMV and spinal cord with subsequent spread to the rostral CNS. These data-driven patterns of αSyn distribution were consistent with established neuropathological staging frameworks. Olfactory-originating subtypes were associated with higher overall brain αSyn burden, whereas brainstem-originating cases showed elevated spinal cord pathology. Early αSyn deposition in the amygdala correlated with higher Alzheimer’s disease co-pathology prevalence, increased APOE ε4 allele frequency, and worse cognitive performance. Conversely, the olfactory-to-brainstem progression trajectory was enriched in cases that showed a higher likelihood of neuropathologically-defined dementia with Lewy bodies. Importantly, the spatial extent of αSyn pathology contributed to dementia independent of Alzheimer and TDP-43 co-pathology.

Current neuropathological staging frameworks for αSyn, including the Braak,^6,7,53,54^ USSLB,^13^ and DLBC,^10,11^ propose different anatomical initiation sites and progression trajectories. These systems conceptualize disease spread as occurring mostly in stereotyped sequential stages, without accounting for the possibility of different spatial subtypes. While the USSLB framework partially acknowledges heterogeneity by distinguishing brainstem-predominant (stage IIa) and limbic-predominant (stage IIb) subtypes, it mostly retains the notion of a consistent sequential staging paradigm with limited individual variability. Recent work proposes the SOC model, distinguishing “body–first” (enteric origin) and “brain–first” (limbic/olfactory origin) phenotypes.^14,15,20^ Employing a data-driven approach, we extend these frameworks to identify different initiation sites and reconstruct subtype-specific progression pathways across the pathological continuum. Our findings reconcile conflicting staging systems: theoretically, DMV-initiated progression aligns with the caudorostral trajectory proposed by Braak, DLBC, and SOC “body-first” models, whereas olfactory-originating subtypes follow the rostrocaudal direction described in the USSLB and SOC “brain-first” pathways. Crucially, we reveal a further heterogeneity within brain-first trajectories such that, where olfactory first subtypes diverge, 50% progressing initially to brainstem regions (similar to USSLB stage IIa) and 50% involving the amygdala earlier (similar to USSLB stage IIb).^10,13^

Ordinal SuStaIn has previously been applied to two hospital-based post-mortem datasets: a large clinical series of 814 individuals with advanced Lewy body disease and a smaller cohort of 173 cases incorporating incidental pathology.^24,25^ While the former benefited from a large sample size, it focused on clinically ascertained cases and lacked peripheral tissue sampling.^25^ Conversely, the latter included extracerebral regions but was limited by hospital-based recruitment and associated comorbidities.^24^ By integrating spinal cord data into a demographically representative population sample, we addressed these gaps in αSyn spatiotemporal modelling. Our findings replicate the results of the large clinical series,^24^ identifying two brain–first trajectories—both originating in the olfactory bulb and diverging toward either the amygdala or brainstem—and one body–first trajectory initiating in the lower brainstem. We also corroborated the specific association between the olfactory–amygdala progression pattern and APOE ɛ4 carriership, Alzheimer’s co-pathology, and severe cognitive decline.^24^ However, we did not observe the sympathetic-predominant subtype reported in the hospital-based extracerebral model.^25^ This discrepancy likely results from anatomical sampling; our study examined the TIML, whereas previous work focused on the sympathetic trunk and cardiac plexus.^25^ As αSyn transmission from the sympathetic trunk to pre-ganglionic neurons in the TIML is hypothesized to be delayed,^55^ the absence of a sympathetic-first trajectory in our cohort, where no isolated TIML pathology occurred, is biologically consistent. Furthermore, while early adrenal involvement characterizes the sympathetic-first subtype,^25^ adrenal pathology in our population emerged only at stage 20. Despite these differences, our data confirm that the subtype originating in the lower brainstem exhibits more extensive extracerebral αSyn burden than those with an olfactory initiation.

The co-occurrence of A*β*, tau, and αSyn pathology is well-documented: 40–60% of Alzheimer’s disease cases exhibit αSyn deposition, predominantly localized to the amygdala and other limbic regions,^56–60^ while 50 – 80% of dementia with Lewy bodies and ∼50% of Parkinson’s disease cases show concomitant Alzheimer’s co-pathology.^61–64^ Consistent with our findings, limbic-predominant synucleinopathy subtypes have been associated with an increased burden of Alzheimer’s co-pathology.^13^ Specifically, in Alzheimer’s disease cases with amygdala-restricted αSyn, inclusions frequently colocalize with tau in the olfactory bulb and amygdala.^56,60^ This colocalization is supported by experimental evidence demonstrating bidirectional cross-seeding between αSyn and tau, which accelerates fibrillogenesis and aggregate propagation.^65–70^ Although αSyn also colocalizes with A*β*, this interaction appears less pronounced.^71,72^ In this study, we demonstrate that Alzheimer’s disease features, including APOE ε4 carriership, advanced tau-related NFT stages, and higher A*β* plaque phases and cortical neuritic plaque densities, are preferentially associated with the progression of αSyn pathology along the olfactory-to-amygdala trajectory. This trajectory likely represents Alzheimer’s disease with amygdala Lewy bodies subtype as a distinct αSyn progression pathway.^9,73,74^ Our results position Alzheimer’s disease co-pathology not merely as a comorbid feature but as an active modulator of αSyn propagation dynamics.

While the olfactory-to-amygdala αSyn trajectory was associated with a higher likelihood of neuropathological Alzheimer’s disease diagnosis, the olfactory-to-brainstem progression more frequently aligned with neuropathologically-defined dementia with Lewy bodies. The SOC model estimates ∼70% of dementia with Lewy bodies cases are “body-first” based on prodromal clinical symptoms.^14,75^ However, our results demonstrate that most individuals with a high likelihood of dementia with Lewy bodies followed the olfactory-to-brainstem trajectory. This discrepancy may stem from our reliance on neuropathological criteria rather than clinical presentation.

In synucleinopathies, αSyn deposition in the neocortex and limbic regions is associated with cognitive decline and dementia.^76,77^ However, the presence of Alzheimer’s co-pathology in people diagnosed with Parkinson’s disease is associated with a higher risk of dementia and shorter intervals from motor symptom onset to dementia.^78,79^ In our study, amygdala-predominant αSyn progression was linked to accelerated cognitive decline and increased prevalence and duration of dementia. Whilst this limbic-predominant subtype was more common in the presence of both Alzheimer’s disease risk factors (APOE ε4 genotype) and Alzheimer’s pathology, we also find that limbic and cortical αSyn pathology alone, in the absence or minimal presence of Alzheimer’s disease pathology, was enough to cause significant cognitive impairment and dementia. This is consistent with recent evidence that Lewy Body and Alzheimer’s disease pathology cause dementia independently.^80^

Several limitations warrant consideration. First, the high dimensionality of regional pathology data relative to cohort size may limit the precision of trajectory inference, raising potential concerns about overfitting. However, the robustness of our model is supported by the concordance of key progression patterns in sensitivity analyses. Second, data availability for peripheral αSyn measures was limited to the adrenal gland and lumbar DRG. Additionally, the lack of systematic sampling may have led to underestimations of pathology outside the CNS. Third, while the population-based design enhances generalizability to community-dwelling older adults, it inherently enriches for early-to-mid stages, limiting insights into end-stage pathology. Furthermore, our cohort consists of individuals aged 85 and older, possibly introducing survivorship bias. Moreover, 81% of our 139 Lewy pathology-positive cases are female, whereas 60% of Parkinson’s disease and 50-60% of diagnosed dementia with Lewy bodies cases are male.^81,82^ Fourth, dementia diagnoses followed DSM-III-R criteria,^83^ which lack the specificity of contemporary frameworks, and data regarding clinical symptoms and diagnoses were not available. Finally, while our cross-sectional model infers temporal progression, validation in longitudinal cohorts with antemortem αSyn-specific biomarkers, and detailed clinical and imaging information may be useful to confirm trajectory accuracy.

Nonetheless, this application of the SuStaIn model to a community-based cohort adds to previous studies in clinically defined samples and supports the theory that αSyn propagation may originate in different areas and employ different routes of propagation, consistent with a body-first and brain-first dichotomy. Close to 45% of these participants over age 85 had evidence of pathogenic αSyn accumulation. The existence of Alzheimer’s disease risk and pathology favours αSyn propagation to the limbic system and neocortex, with consequent dementia. Crucially, our findings establish corticolimbic αSyn as an independent pathogenic contributor to dementia, separate from the influence of Alzheimer’s disease and TDP-43 comorbidities.

## Data availability

The data that support the findings of this study are available from the corresponding author upon reasonable request, subject to relevant approvals.

## Supplementary material

Supplementary material is available on medRxiv.

