## Supplementary Materials for "Alpha-synuclein propagation trajectories in a population-based postmortem cohort"

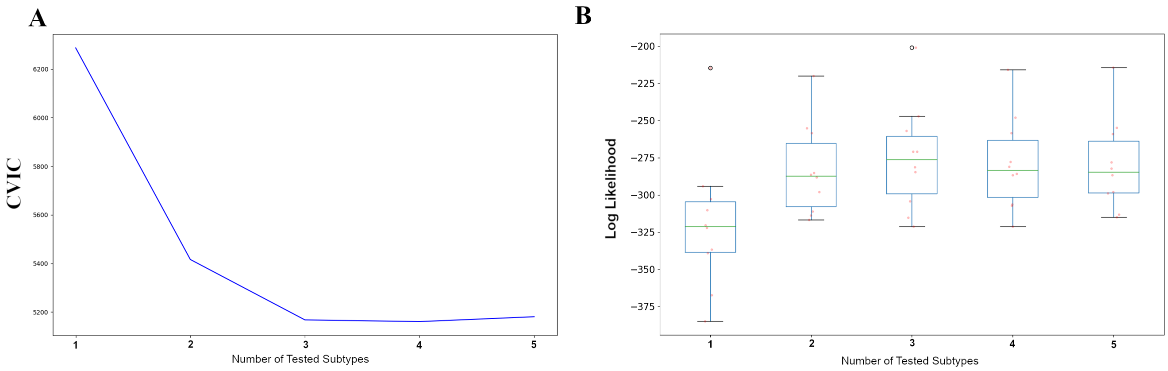


**Supplementary Figure 1. Determination of the optimal number of SuStaIn subtypes**. The SuStaIn model was fitted with 1 to 5 subtypes. (**A**) Cross-validation information criterion (CVIC) and (**B**) log-likelihood values indicate that a three-subtype solution provides the optimal model fit.

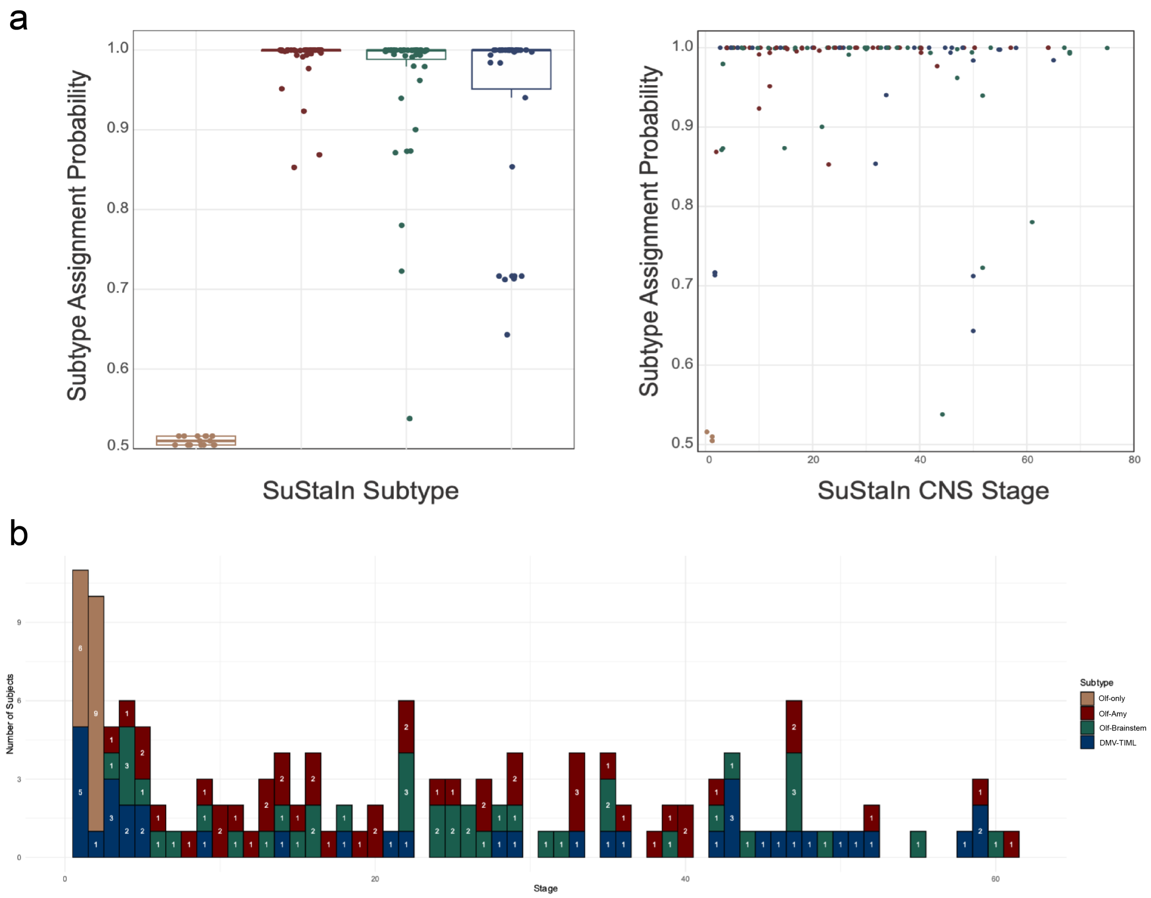


**Supplementary Figure 2. SuStaIn subtype assignment probabilities and stage distributions.** (**A**) Probabilities of assignment to each SuStaIn subtype across for each subtype (left) and along SuStaIn stages (right). (**B**) Distribution of SuStaIn stages, shown for each subtype: Olf-only (orange), Olf-Amy (red), Olf-Brainstem (green), and DMV-TIML (blue). Olf-Amy, olfactory-amygdala; Olf-Brainstem, olfactory-brainstem; Olf-only, olfactory-only, DMV-TIML, dorsal motor nucleus of the vagus–thoracic intermediolateral nucleus; SuStaIn, Subtype and Stage Inference.

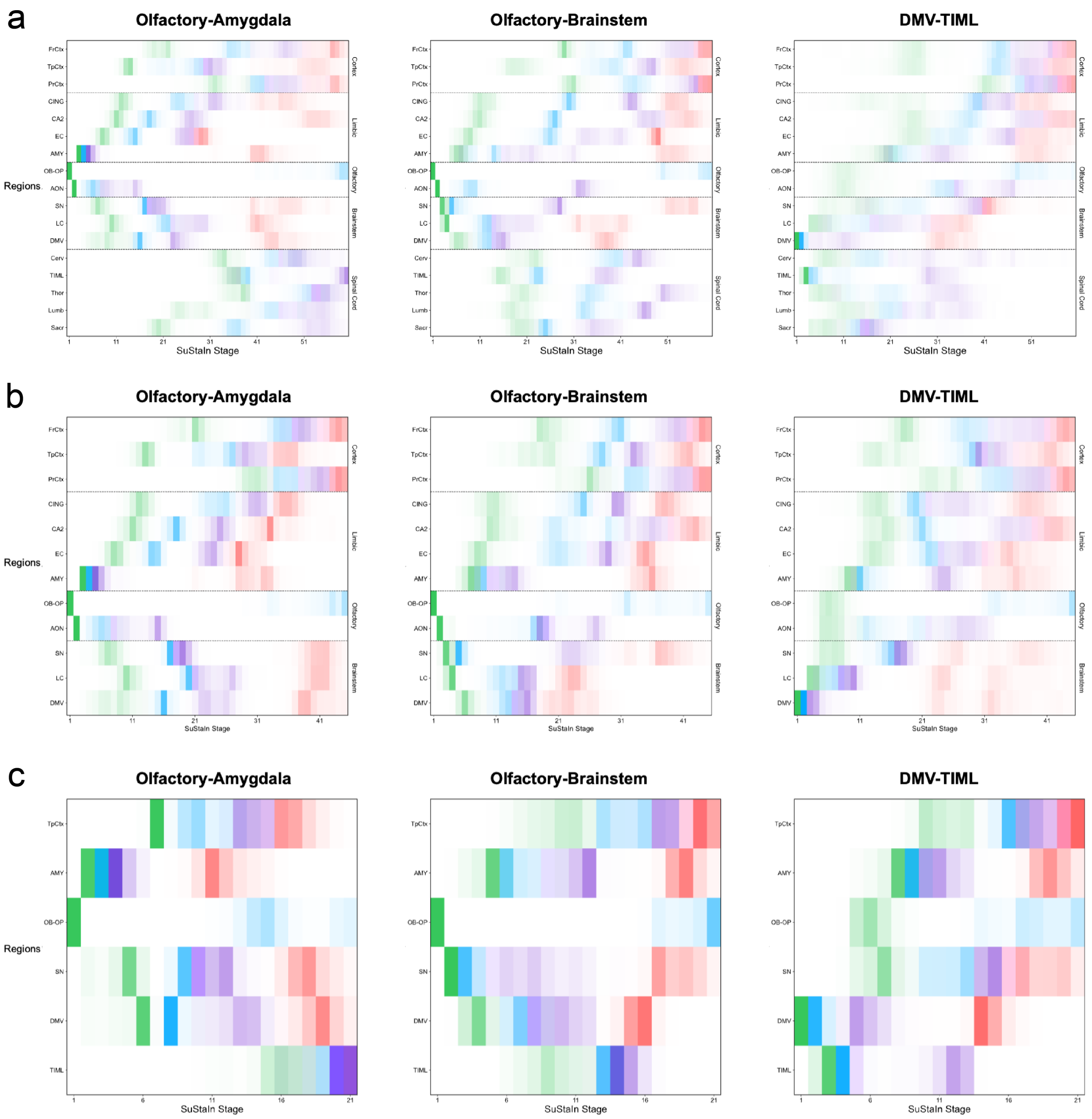


**Supplementary Figure 3. Sensitivity analyses of SuStaIn-inferred αSyn trajectories.** Positional variance diagrams depict the stability of subtype trajectories under different data conditions: (**A**) Analysis restricted to individuals with complete data, (**B**) analysis using exclusively brain regions, and (**C**) analysis using a reduced feature set comprising six regions (TpCtx, AMY, OB-OP, SN, DMV, and TIML). Darker shading indicates higher certainty of regional involvement, color hue represents pathology severity (green = mild, blue = moderate, purple = severe, red = very severe), and white boxes indicate no transitions in severity. ADR, adrenal gland; AON, anterior olfactory nucleus; AMY, amygdala; CA2, cornu ammonis 2; Cerv, cervical spinal cord; CING, cingulate gyrus; DMV, dorsal motor nucleus of the vagus; DRG, dorsal root ganglion; EC, entorhinal cortex; FrCtx, frontal cortex; LC, locus coeruleus; Lumb, lumbar spinal cord; OB-OP, peripheral olfactory bulb/peduncle; Olf, olfactory; PrCtx, parietal cortex; SN, substantia nigra; Sacr, sacral spinal cord; Thor, thoracic spinal cord; TIML, thoracic intermediolateral column; TpCtx, temporal cortex; SuStaIn, Subtype and Stage Inference.

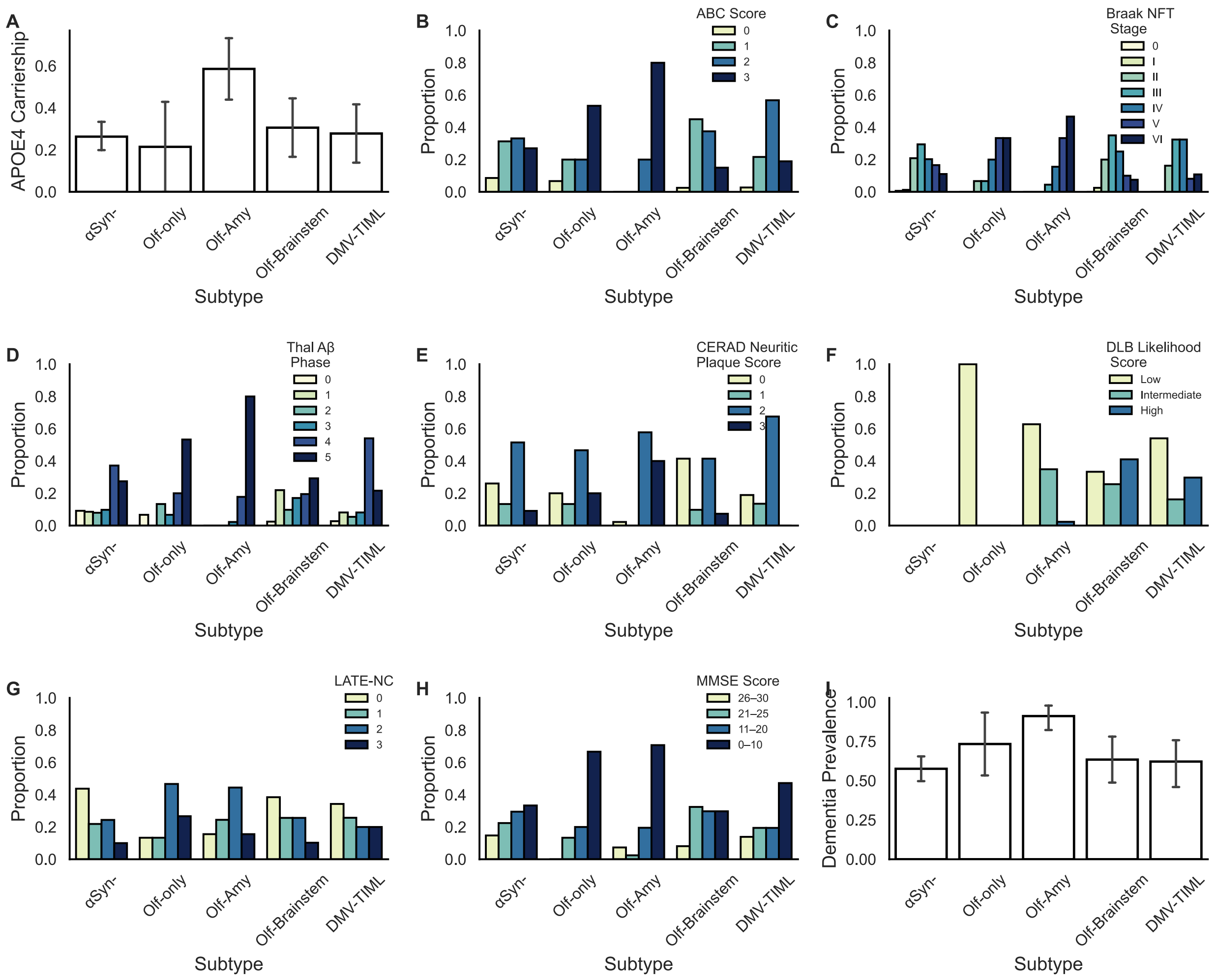

**Supplementary Figure 4. Comparisons of αSyn-, Olf-only, and SuStaIn subtypes across clinical, neuropathological, and genetic metrics.** (**A**) APOE carrier status, (**B**) ABC score, (**C**) Braak NFT stage, (**D**) Thal A$\beta$ phase, (**E**) CERAD neuritic plaque score, (**F**) DLB likelihood score, (**G**) LATE-NC stage, (**H**) MMSE score, and (**I**) dementia prevalence. A$\beta$, amyloid-$\beta$; APOE, apolipoprotein E; CERAD, Consortium to Establish a Registry for Alzheimer's Disease; DLB, dementia with Lewy bodies; DMV-TIML, dorsal motor nucleus of the vagus–thoracic intermediolateral nucleus; LATE-NC, limbic-predominant age-related TDP-43 encephalopathy; MMSE, Mini-Mental State Examination; NFT, neurofibrillary tangle; Olf-Amy, olfactory-amygdala; Olf-Brainstem, olfactory-brainstem; SuStaIn, Subtype and Stage Inference.

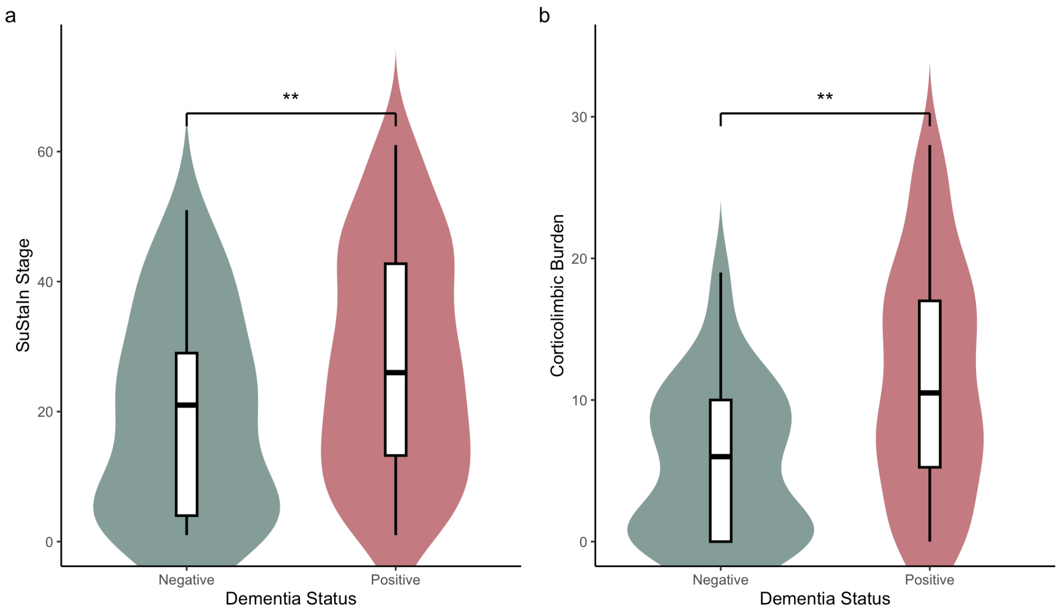


**Supplementary Figure 5. SuStaIn stage (A) and corticolimbic αSyn burden (B) in αSyn+ demented versus non-demented individuals with no or low Alzheimer's disease neuropathological change.** Statistical significance is indicated as $*p<0.05$, and $**p<0.01$.

**Supplementary Table 1. Anatomical regions and scoring criteria for α-synuclein pathology**

| Anatomical Division | Region | Specific Sub-regions / Levels | Scoring Scale |
|---|---|---|---|
| **Brain (Neocortex)** | Frontal Cortex | Medial and superior gyrus | 0–4 |
| | Temporal Cortex | Medial and superior gyrus | 0–4 |
| | Parietal Cortex | Inferior and superior lobule | 0–4 |
| **Brain (Limbic)** | Cingulate Cortex | Cingulate gyrus | 0–4 |
| | Hippocampus | Cornu ammonis 2 | 0–4 |
| | | Entorhinal cortex | 0–4 |
| | Amygdala | | 0–4 |
| **Brain (Brainstem)** | Midbrain | Substantia nigra (SN) | 0–4 |
| | Pons | Locus coeruleus (LC) | 0–4 |
| | Medulla Oblongata | Dorsal motor nucleus of the vagus (DMV) | 0–4 |
| **Brain (Olfactory)** | Olfactory Bulb | Peripheral olfactory bulb & peduncle (OB-OP) | 0–3 |
| | | Anterior olfactory nucleus (AON) | 0–3 |
| **Spinal Cord** | Cervical | Levels C6–C7 (Dorsal & Ventral horns) | 0–3 |
| | Thoracic | Levels T3–T4 (Dorsal & Ventral horns) | 0–3 |
| | | Thoracic Intermediolateral Column (TIML) | 0–3 |
| | Lumbar | Levels L3–L4 (Dorsal & Ventral horns) | 0–3 |
| | Sacral | Levels S1–S2 (Dorsal & Ventral horns) | 0–3 |
| **Peripheral** | Dorsal Root Ganglia | Lumbar level | 0–1 |
| | Adrenal Gland | Adrenal medulla | 0–1 |

Note: α-Synuclein pathology was assessed using immunohistochemical staining with a mouse monoclonal antibody (clone 5G4, dilution 1:1000). 0–4 scale: Scored according to the consensus guidelines of the third Dementia with Lewy Bodies Consortium. 0 = no Lewy bodies (LB) or Lewy neurites (LN); 1 = mild (1 LB or sparse LNs); 2 = moderate (≥2 LBs or scattered LNs); 3 = severe (≥4 LBs or numerous LNs); 4 = very severe (numerous LBs and LNs). 0–3 scale: Modified scale where severe and very severe categories were combined to preserve statistical power. 0 = absent; 1 = mild; 2 = moderate; 3 = severe/very severe. 0–1 scale: Dichotomous scoring. 0 = absent; 1 = present.

**Supplementary Table 2. Conversion of neuropathological density scores to probability scores for regions with scores from 0 to 4 (neocortex, limbic, and brainstem regions)**

| | | Score Probability | | | | |
|---|---|---|---|---|---|---|
| | | 0 | 1 | 2 | 3 | 4 |
| Neuropathological Density Score | 0 | 0.881 | 0.119 | $2.95 \times 10^{-4}$ | $1.34 \times 10^{-8}$ | $1.12 \times 10^{-14}$ |
| | 1 | 0.107 | 0.787 | 0.107 | $2.64 \times 10^{-4}$ | $1.20 \times 10^{-8}$ |
| | 2 | $2.64 \times 10^{-4}$ | 0.107 | 0.787 | 0.107 | $2.64 \times 10^{-4}$ |
| | 3 | $1.20 \times 10^{-8}$ | $2.64 \times 10^{-4}$ | 0.107 | 0.787 | 0.107 |
| | 4 | $1.12 \times 10^{-14}$ | $1.34 \times 10^{-8}$ | $2.95 \times 10^{-4}$ | 0.119 | 0.881 |

**Supplementary Table 3. Conversion of neuropathological density scores to probability scores for regions with scores from 0 to 3 (olfactory and spinal cord regions)**

| | | Score Probability | | | |
|---|---|---|---|---|---|
| | | 0 | 1 | 2 | 3 |
| Neuropathological Density Score | 0 | 0.881 | 0.119 | $2.95 \times 10^{-4}$ | $1.34 \times 10^{-8}$ |
| | 1 | 0.107 | 0.787 | 0.107 | $2.64 \times 10^{-4}$ |
| | 2 | $2.64 \times 10^{-4}$ | 0.107 | 0.787 | 0.107 |
| | 3 | $1.34 \times 10^{-8}$ | $2.95 \times 10^{-4}$ | 0.119 | 0.881 |

**Supplementary Table 4. Conversion of neuropathological density scores to probability scores for regions with dichotomous scores (adrenal gland and lumbar DRG)**

| | | Score Probability | |
|---|---|---|---|
| | | 0 | 1 |
| Neuropathological Density Score | 0 | 0.881 | 0.119 |
| | 1 | 0.119 | 0.881 |

**Supplementary Table 5. Demographic, clinical, and neuropathological characteristics of αSyn-, Olfactory-only, and SuStaIn Subtypes.**

| | | **Olf-Amy (*n*=45)** | **Olf-Brainstem (*n*=41)** | **DMV-TIML (*n*=37)** | **αSyn- (*n*=165)** | **Olf-only (*n*=15)** |
|---|---|---|---|---|---|---|
| **Age at death** (years), mean±SD | | 92.74 ± 4.06 | 91.57 ± 3.44 | 93.10 ± 4.73 | 92.29 ± 3.46 | 92.73 ± 4.00 |
| **Sex** (Female), n(%) | | 38 (84) | 30 (73) | 30 (81) | 139 (84) | 14 (93) |
| **APOE-ε4 carriers**, n(%) | | 24 (59) | 11 (31) | 10 (28) | 41 (26) | 3 (21) |
| **Dementia**, n(%) | | 41 (91) | 26 (63) | 23 (62) | 95 (58) | 11 (73) |
| **Dementia duration** (years), mean±SD | | 6.8 ± 3.8 | 3.1 ± 2.1 | 5.2 ± 3.7 | 5.0 ± 3.7 | 6.8 ± 4.5 |
| **MMSE score, mean±SD** | | 6.4 ± 8.9 | 15.7 ± 8.6 | 13.1 ± 9.9 | 15.1 ± 8.8 | 8.6 ± 8.7 |
| **LATE-NC stage** n(%) | 0 | 7 (16) | 15 (39) | 12 (34) | 70 (44) | 2 (13) |
| | 1 | 11 (24) | 10 (26) | 9 (26) | 35 (22) | 2(13) |
| | 2 | 20 (44) | 10 (26) | 7 (20) | 39 (24) | 7 (47) |
| | 3 | 7 (16) | 4 (10) | 7 (20) | 16 (10) | 4 (27) |
| **Thal phase**, n(%) | 0-2 | 0 (0) | 14 (34) | 6 (16) | 42 (26) | 3 (20.0) |
| | 3-4 | 9 (20) | 15 (37) | 23 (62) | 77 (47) | 4 (27) |
| | 5 | 36 (80) | 12 (29) | 8 (22) | 45 (27) | 8 (53) |
| **Braak NFT stage**, n(%) | 0-II | 0 (0) | 9 (23) | 6 (16) | 37 (23) | 1 (7) |
| | III-IV | 9 (20) | 24 (60) | 24 (65) | 81 (50) | 4 (27) |
| | V-VI | 36 (80) | 7 (18) | 7 (19) | 45 (28) | 10 (67) |
| **CERAD score**, n(%) | None-Sparse | 1 (2) | 21 (51) | 12 (32) | 65 (39) | 5 (33) |
| | Moderate | 26 (58) | 17 (42) | 25 (68) | 85 (52) | 7 (47) |
| | Frequent | 18 (40) | 3 (7) | 0 (0) | 15 (9) | 3 (20) |
| **ABC score**, n(%) | Not/Low | 0 (0) | 19 (48) | 9 (24) | 65 (40) | 4 (27) |
| | Intermediate | 9 (20) | 15 (38) | 21 (57) | 54 (33) | 3 (20) |
| | High | 36 (80) | 6 (15) | 7 (19) | 44 (27) | 8 (53) |

Subtype characteristics are presented as n (%) unless otherwise indicated. αSyn, α-synuclein; CERAD, Consortium to Establish a Registry for Alzheimer's Disease; DMV-TIML, dorsal motor nucleus of the vagus–thoracic intermediolateral nucleus; LATE-NC, limbic-predominant age-related TDP-43 encephalopathy; MMSE, Mini-Mental State Examination; NFT, neurofibrillary tangle; Olf-Amy, olfactory-amygdala; Olf-Brainstem, olfactory-brainstem; Olf-only: Olfactory-only SuStaIn, Subtype and Stage Inference.

**Supplementary Table 6. Regression analyses examining pathological and clinical predictors of dementia and cognitive performance.**

| | | **Dementia (Model 1),** OR (95%CI) | **Dementia (Model 2),** OR (95%CI) | **MMSE (Model 3),** *β* (95%CI) | **MMSE (Model 4),** *β* (95%CI) |
|---|---|---|---|---|---|
| **Intercept** | | 0.07 (0.00, 68.04) | 0.06 (0.00, 61.53) | 38.69 (14.27, 63.11) ** | 39.73 (15.38, 64.08) ** |
| **Age at death** | | 1.02 (0.94, 1.09) | 1.02 (0.94, 1.10) | -0.16 (-0.42, 0.09) | -0.18 (-0.44, 0.08) |
| **Sex (Female)** | | 1.68 (0.82, 3.46) | 1.68 (0.82, 3.43) | -5.18 (-7.77, -2.57) *** | -5.15 (-7.73, -2.56) *** |
| **ABC score** (0/1 as reference) | Score 2 | 1.60 (0.87, 2.95) | 1.57 (0.85, 2.90) | -2.49 (-4.86, -0.10) * | -2.44 (-4.81, -0.06) * |
| | Score 3 | 7.98 (3.80, 17.87) *** | 7.41 (3.51, 16.65) *** | -7.13 (-9.62, -4.63) *** | -6.84 (-9.35, -4.34) *** |
| **LATE-NC stage** (0/1 as reference) | Stage 2 | 2.09 (1.09, 4.13)* | 2.05 (1.06, 4.08)* | -4.06 (-6.40, -1.70)*** | -3.96 (-6.31, -1.62)*** |
| | Stage 3 | 5.53 (2.12, 17.35)** | 5.70 (2.17, 17.95)** | -6.73 (-9.67, -3.78)*** | -6.65 (-9.58, -3.72)*** |
| **αSyn SuStaIn stage** | | 1.03 (1.01, 1.05) ** | - | -0.06 (-0.12, - 0.01) * | - |
| **αSyn Corticolimbic burden** | | - | 1.09 (1.03, 1.15) ** | - | -0.18 (-0.33, - 0.03) * |

For the dependent variable dementia, Models 1 and 2 were estimated using logistic regression, with results reported as odds ratios (ORs) and 95% confidence intervals (CIs). Model 1 includes the αSyn SuStaIn stage, whereas Model 2 includes the αSyn corticolimbic burden. For the dependent variable MMSE, Models 3 and 4 were estimated using linear regression and report unstandardized coefficients (*β*) with 95% CIs. Statistical significance is denoted as *p<0.05, **p<0.01, and ***p<0.001. αSyn, α-synuclein; LATE-NC, limbic-predominant age-related TDP-43 encephalopathy; MMSE, Mini-Mental State Examination; SuStaIn, Subtype and Stage Inference.
